# Attitudes and Experiences Surrounding Female Genital Mutilation/Cutting in the United States: A Scoping Review

**DOI:** 10.1101/2022.03.25.22272941

**Authors:** Ghenet Besera, Howard Goldberg, Ekwutosi M Okoroh, Margaret Christine Snead, Crista E Johnson-Agbakwu, Mary M Goodwin

**Author notes:** Corresponding Author: Margaret Christine Snead, Division of Reproductive Health, Centers for Disease Control and Prevention 4770 Buford Hwy, NE, Atlanta, GA 30341.

## Abstract

In recent decades, growing migration to the United States from countries where female genital mutilation/cutting (FGM/C) is widely practiced has caused a rise in the number of women and girls in the United States who could have potentially experienced FGM/C. A scoping review was conducted to identify research and gaps in literature about FGM/C–related attitudes and experiences among individuals from FGM/C–practicing countries living in the United States. This scoping review identified 40 articles meeting inclusion criteria. The findings of this review suggest that both women and men from FGM/C-practicing countries living in the United States generally oppose FGM/C, and that women with FGM/C have significant physical and mental health needs and have found US healthcare providers to lack understanding of FGM/C. Future research can improve measurement of FGM/C by applying a health equity lens and taking into account the sociocultural influences on FGM/C–related attitudes and experiences.

## INTRODUCTION

Female genital mutilation/cutting (FGM/C) is a health and human rights concern defined by the World Health Organization (WHO) as “all procedures involving partial or total removal of the external female genitalia or other injury to the female genital organs for non-medical reasons” [1]. WHO classifies FGM/C into four types (Table 1). FGM/C can have health consequences that are both immediate (e.g., pain, bleeding, swelling) [2] and long-term (e.g., obstetric complications, painful intercourse, psychological problems) [2, 3]. FGM/C has deep cultural and social roots, and among groups who practice FGM/C, reasons given for carrying out the practice include traditional beliefs, attitudes, and practices such as rites of passage to adulthood, social acceptance, religion, hygiene, curtailing girls’ sexuality, preventing promiscuity, marriageability, and honor [4, 5]. It is estimated that at least 200 million women and girls have experienced FGM/C globally, with the practice concentrated in 30 countries in Africa, Asia, and the Middle East [6]. Previous studies have found FGM/C among women and girls who migrated from FGM/C-practicing countries to other countries, including in Australia, Europe, and North America [7–9].

**Table 1.** World Health Organization Female Genital Mutilation/Cutting Types.

|  |  |
| --- | --- |
| **Type I** | “Partial or total removal of the clitoris and/or the prepuce (clitoridectomy)” |
| **Type II** | “Partial or total removal of the clitoris and the labia minora, with or without excision of the labia majora (excision)” |
| **Type III** | “Narrowing of the vaginal orifice with creation of a covering seal by cutting and appositioning the labia minora and/or the labia majora, with or without excision of the clitoris (infibulation)” |
| **Type IV** | “All other harmful procedures to the female genitalia for non-medical purposes, for example: pricking, piercing, incising, scraping and cauterization” |
| *Source: World Health Organization, 2008, p. 4 [1]* |  |

In the United States, FGM/C is illegal, as is the act of transporting an individual from the United States to other countries for FGM/C (known as “vacation cutting”) [10–12]. A 2016 study estimated that as many as 513,000 women and girls in the United States could have experienced or be at risk of FGM/C, a threefold increase from a 1990 estimation using the same methodology [13, 14]. These estimates applied the prevalence of FGM/C in countries of origin to US populations of women from countries where FGM/C is practiced [13, 14]. Since both estimates used indirect methods, two important limitations must be noted. One is the inability to differentiate the number of women who may have experienced FGM/C before emigration from those who could be at risk in the future [13]. The second is that estimates about FGM/C in the United States carry the assumption that people from countries where FGM/C is common will maintain the same attitudes and practices after migration. In recent decades, an increase in migration to the United States from countries where FGM/C is prevalent has increased the potential number of females who experienced or may be at risk of experiencing FGM/C [13].

Although the exact number of women and girls living with FGM/C in the United States is unknown [13, 14], increasing attention has been directed toward local and national efforts aimed at FGM/C prevention and providing support and care to women and girls who have undergone FGM/C [15, 16]. To further address this public health issue, it is important to not only quantify the number of people affected but to understand their perspectives on the practice and how they may change after migration. To our knowledge, no published studies synthesize the literature on FGM/C in the United States. To fill this gap, we conducted a scoping review of the existing literature on attitudes and experiences related to FGM/C among US residents from FGM/C– practicing countries to identify and consolidate existing research on the topic and assess gaps to better guide future research and programmatic efforts addressing FGM/C in the United States.

## METHODS

This scoping review was guided by the framework developed by Arksey and O’Malley [17]. The guiding research question is, what evidence is available about the attitudes and experiences surrounding FGM/C among US-resident individuals from FGM/C–practicing countries? The search included the key words or Medical Subject Headings (MeSH) terms “Female Circumcision,” “Female Genital Mutilation,” “Female Genital Cutting,” and “United States.” A health sciences librarian helped search Medline (OVID), Embase (OVID), PubMed, and SCOPUS for relevant literature published in English through April 2021 (with no limitation on the earliest publication date). We conducted a targeted gray literature search of organizational websites, Google, Google Scholar, and ProQuest Dissertations. We also reviewed references of relevant articles and identified those that potentially met inclusion criteria. Once we collected articles, we exported all records to Endnote to check for duplicates. We then imported records to Covidence, a web-based software that facilitates reference screening and data extraction [18].

The next stage of the scoping review involved selecting relevant articles through a two-level screening process to determine if they met the inclusion criteria, which included:

- Data collected directly from women, men, or girls from FGM/C-practicing countries living in the United States.

○ If studies included other populations (e.g., study participants not living in the United States, medical professionals), they must have disaggregated findings for women, men, or girls living in the United States. Studies that only included healthcare providers’ attitudes and experiences or perspectives of non-US-based individuals were excluded.
○ Studies that were non-data driven (e.g., commentaries, literature reviews) or did not present data collected directly from the populations of interest did not meet the inclusion criteria.
- Studies assessing either attitudes or experiences related to FGM/C.
- Availability of the full text of the article.

There were no other study design or methodological restrictions.

Study authors (G.B and H.G) independently reviewed each article’s title and abstract to find articles that potentially met the inclusion criteria. At the second screening level, they independently reviewed the full articles identified during the first review. Both reviewers assessed and had consensus on all studies included in the review. Discrepancies were reconciled during meetings among study authors (G.B and H.G) to ensure consistency in the review strategy and with inclusion/exclusion decisions.

The study selection process and results are presented in a Preferred Reporting Items for Systematic Reviews and Meta-Analyses (PRISMA) flow diagram (Figure 1) [19]. Once articles meeting inclusion criteria were identified, the two reviewers extracted study information and conducted a quality appraisal of all studies using the National Heart, Lung, and Blood Institute Quality Assessment Tool for Observational Cohort and Cross-Sectional Studies for quantitative studies [20], Critical Appraisal Skills Program checklist for qualitative studies [21], and Mixed Methods Appraisal Tool for mixed methods studies [22]. Reviewers gave an overall quality rating to each study and resolved discrepancies in any decisions through discussion. Overall, 26 studies were rated as “good/excellent,” and the remaining 14 were rated as “fair.” No studies were rated as “poor.” Since this was a review of published studies, institutional review board review was not necessary.

**Figure 1.**
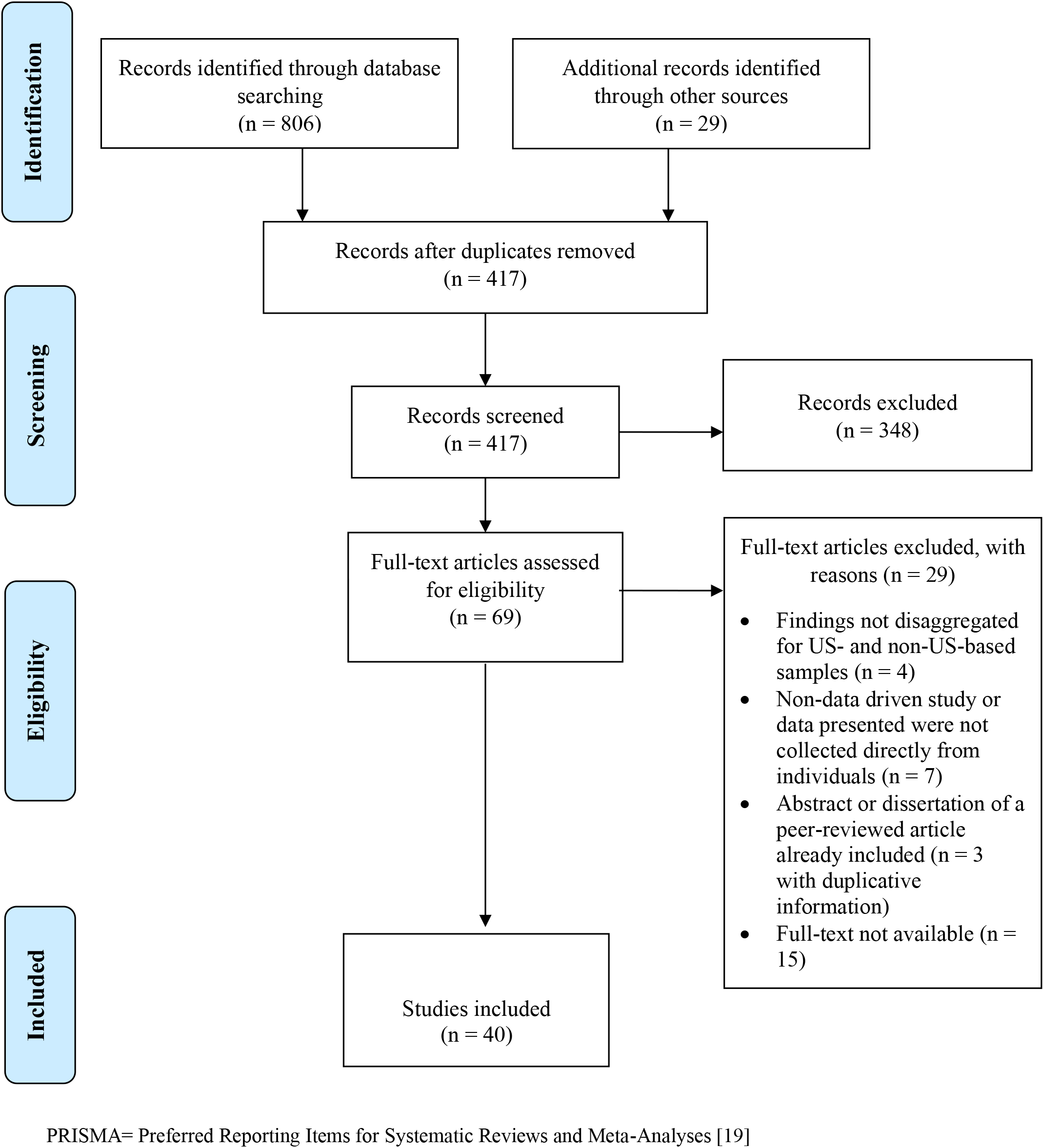
PRISMA Flow Diagram of Article Selection.

## RESULTS

### Study Selection

After we removed duplicates, a total of 417 articles remained on the original list, including 388 records identified through database searches and 29 identified through other sources. After title and abstract screening, 69 studies remained. During full-text screening, we excluded 29 articles (Figure 1). The final 40 articles consisted of 32 peer-reviewed articles, 7 dissertations, and one evaluation report (Table 2). Scoping review authors grouped findings into major themes and categories; results are organized and presented accordingly in the sections below.

**Table 2.**
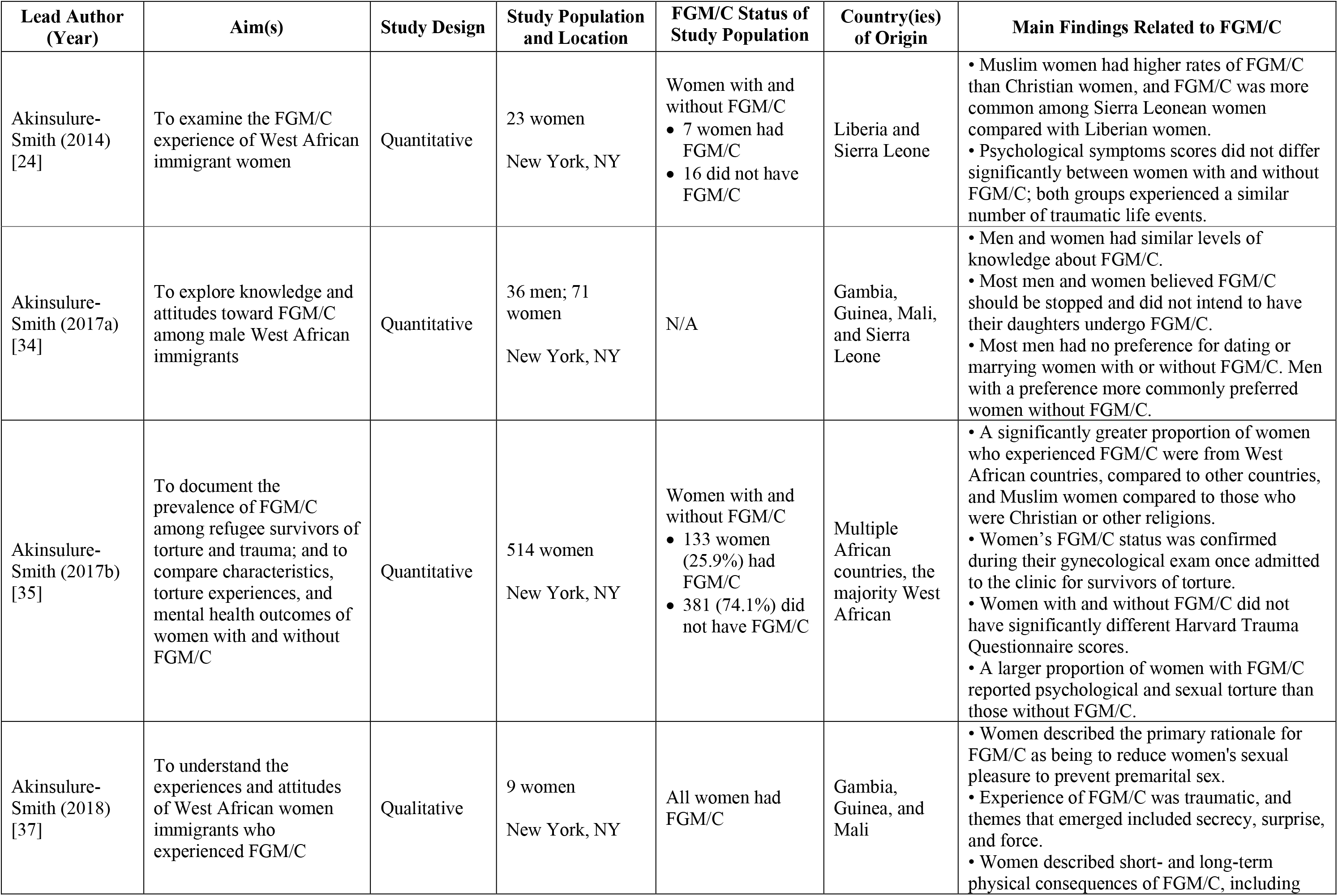
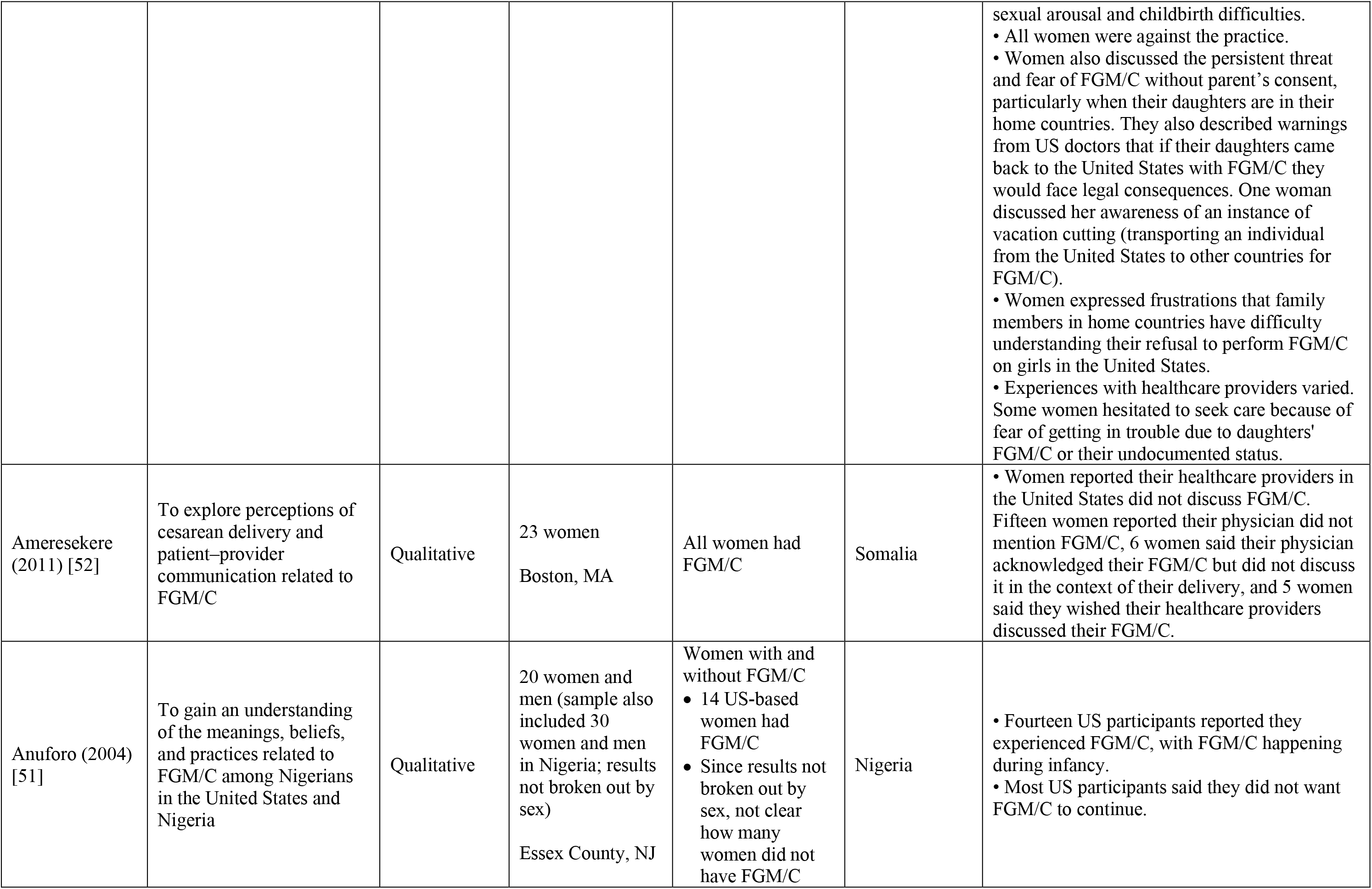
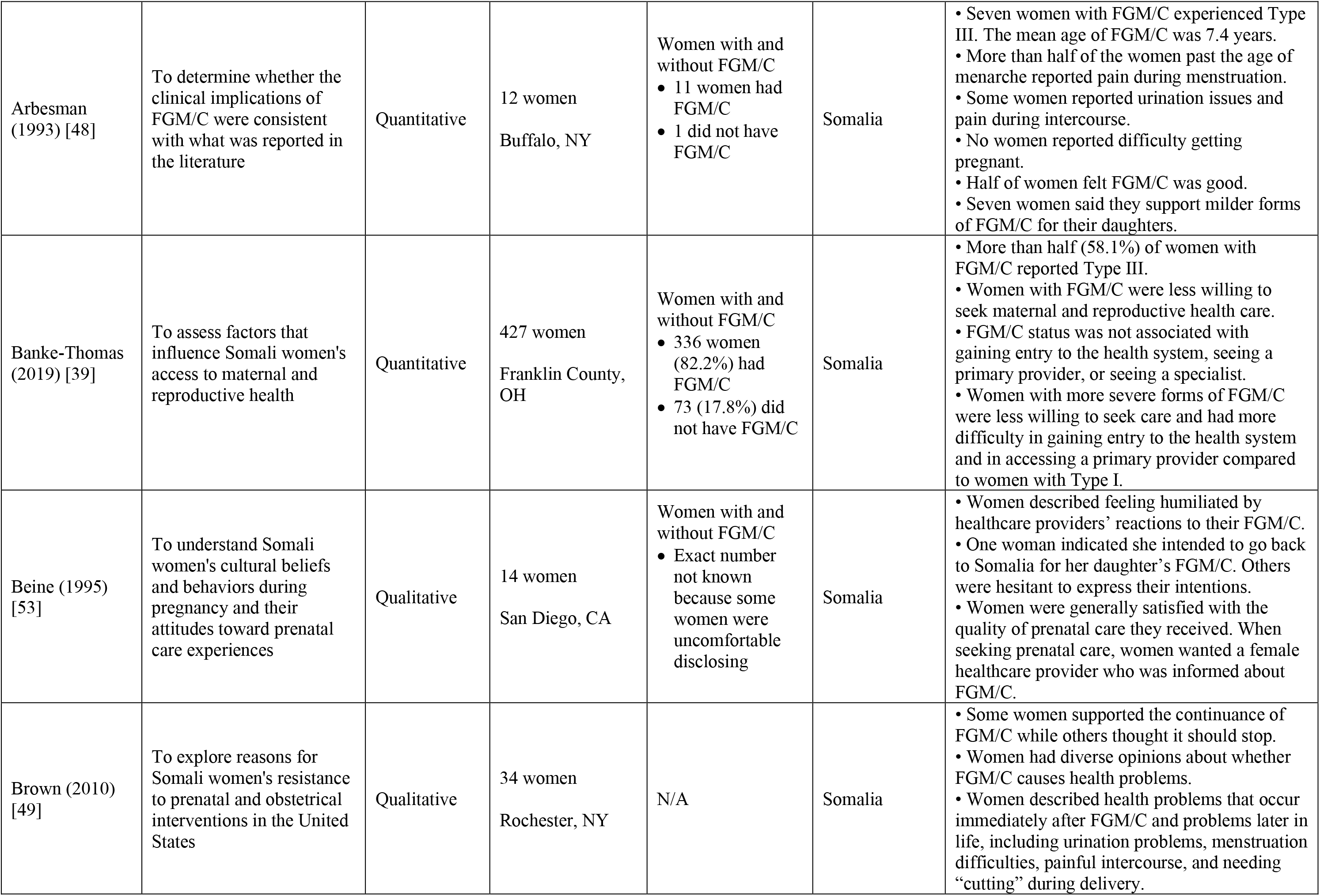
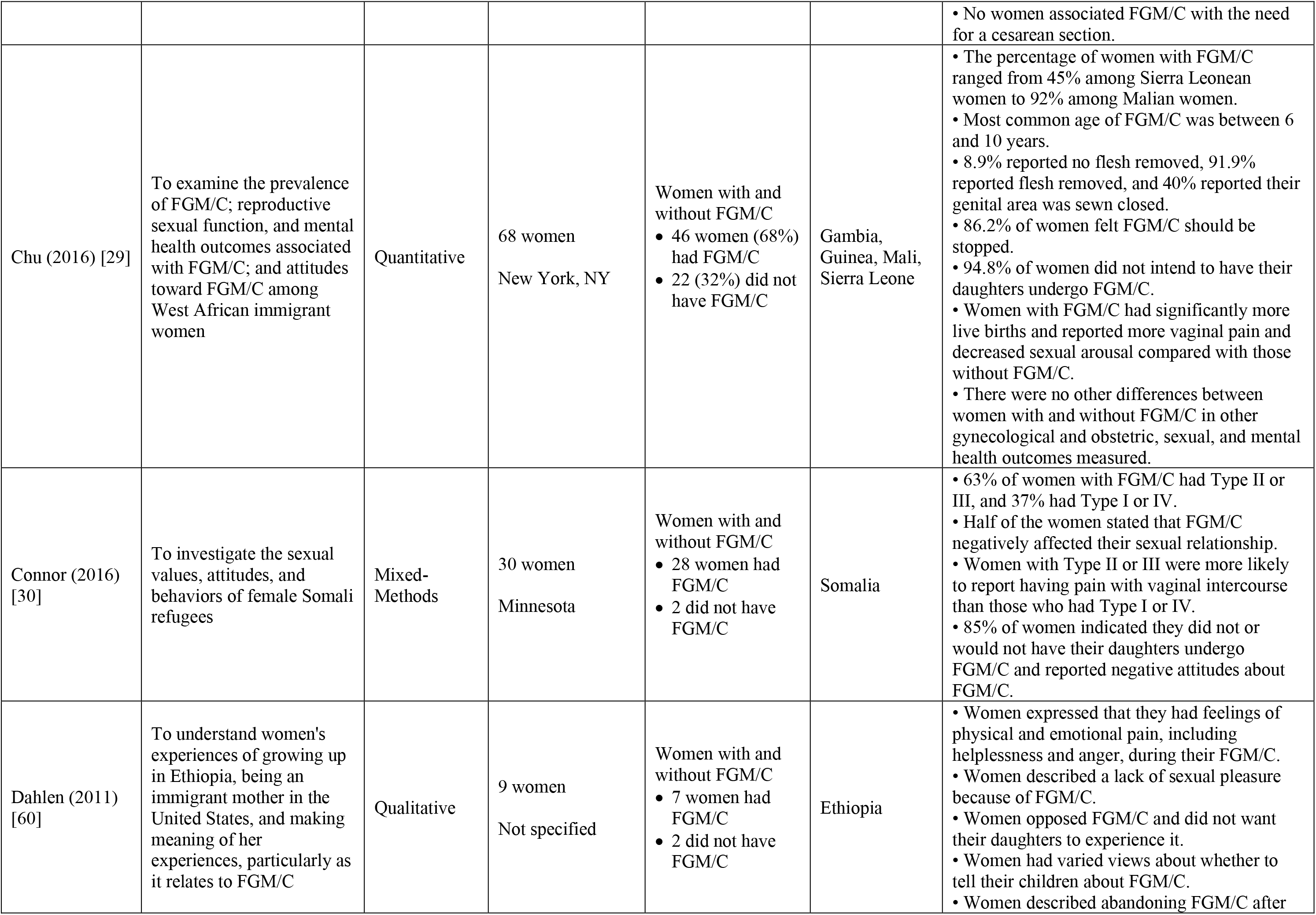
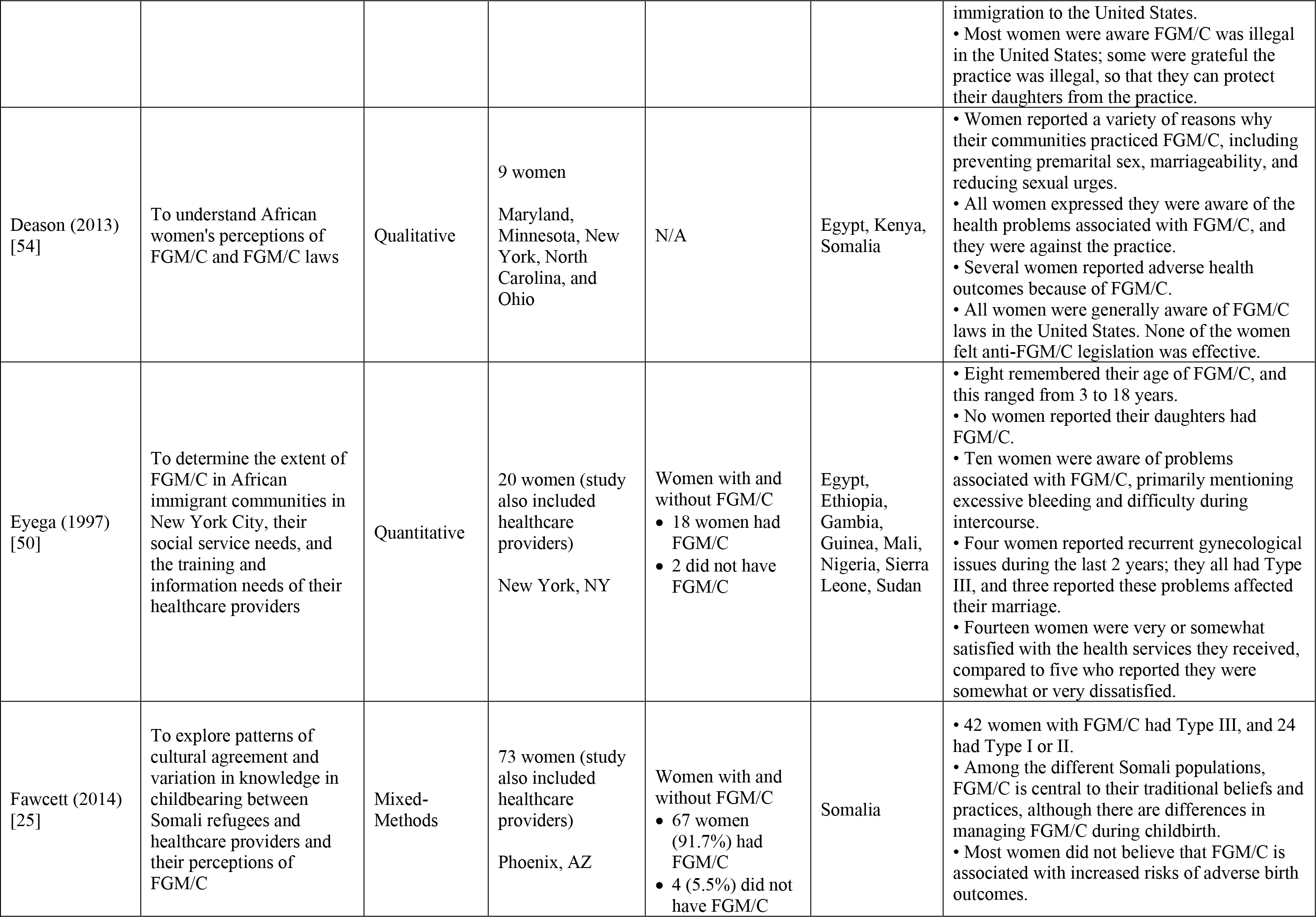
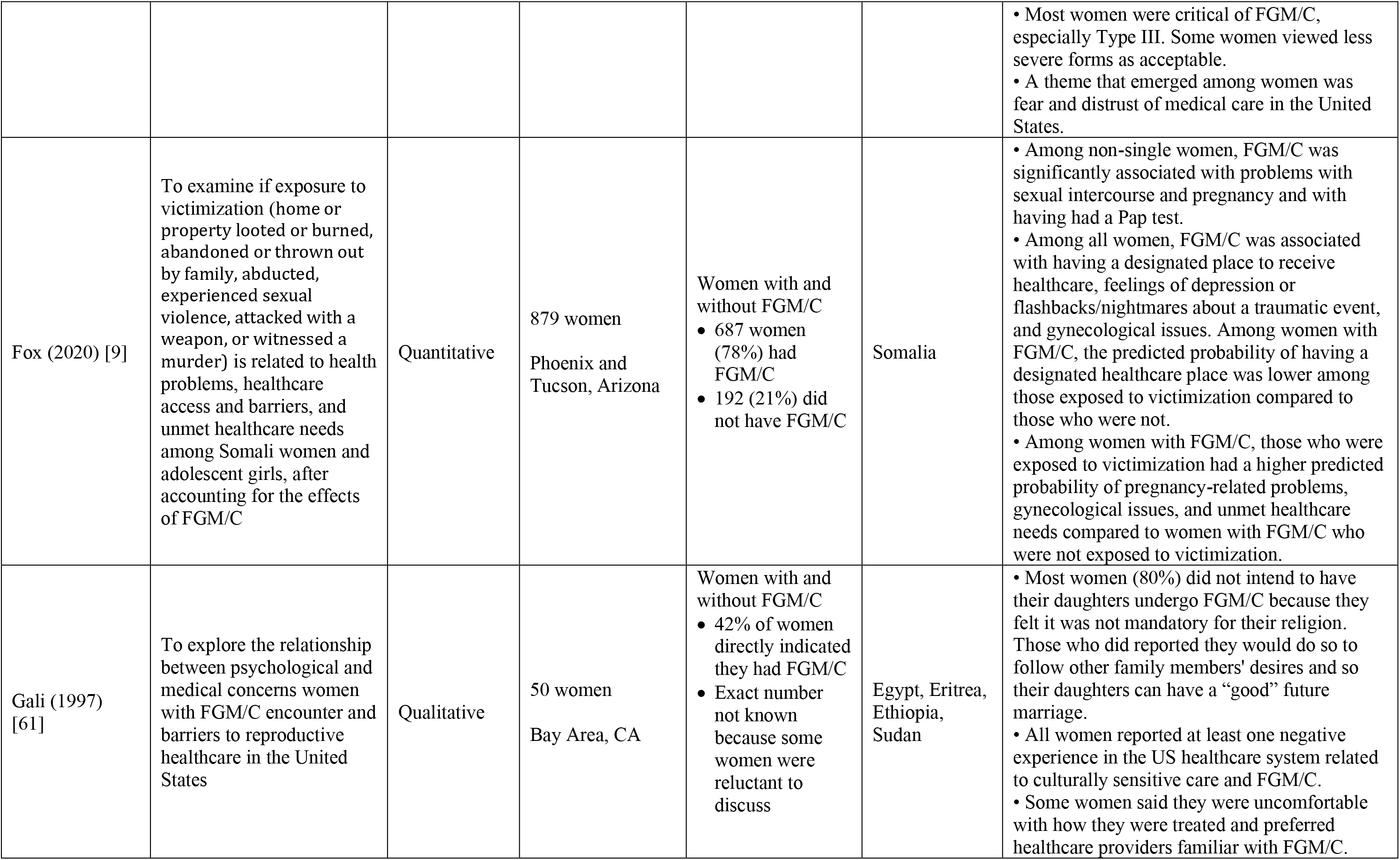
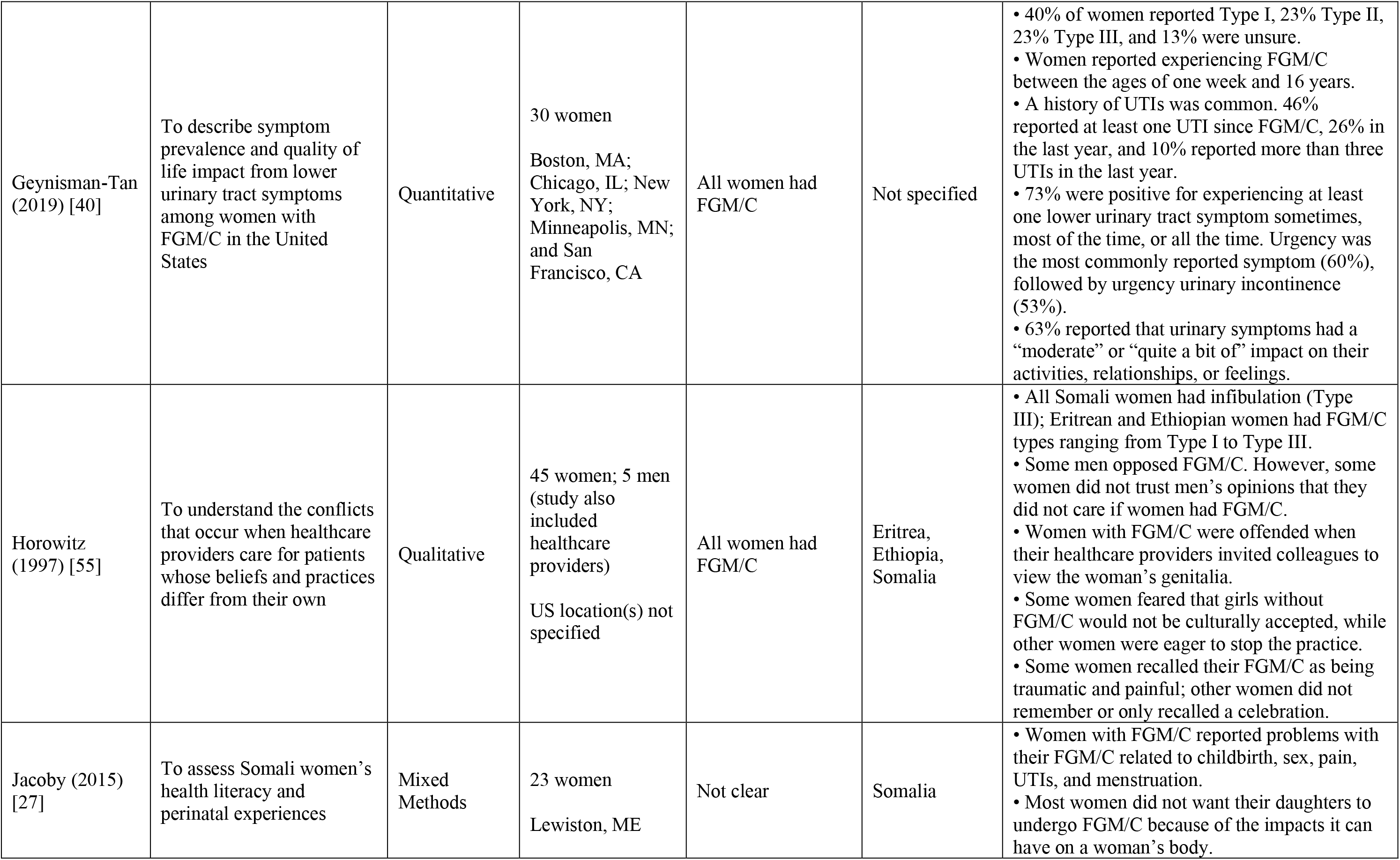
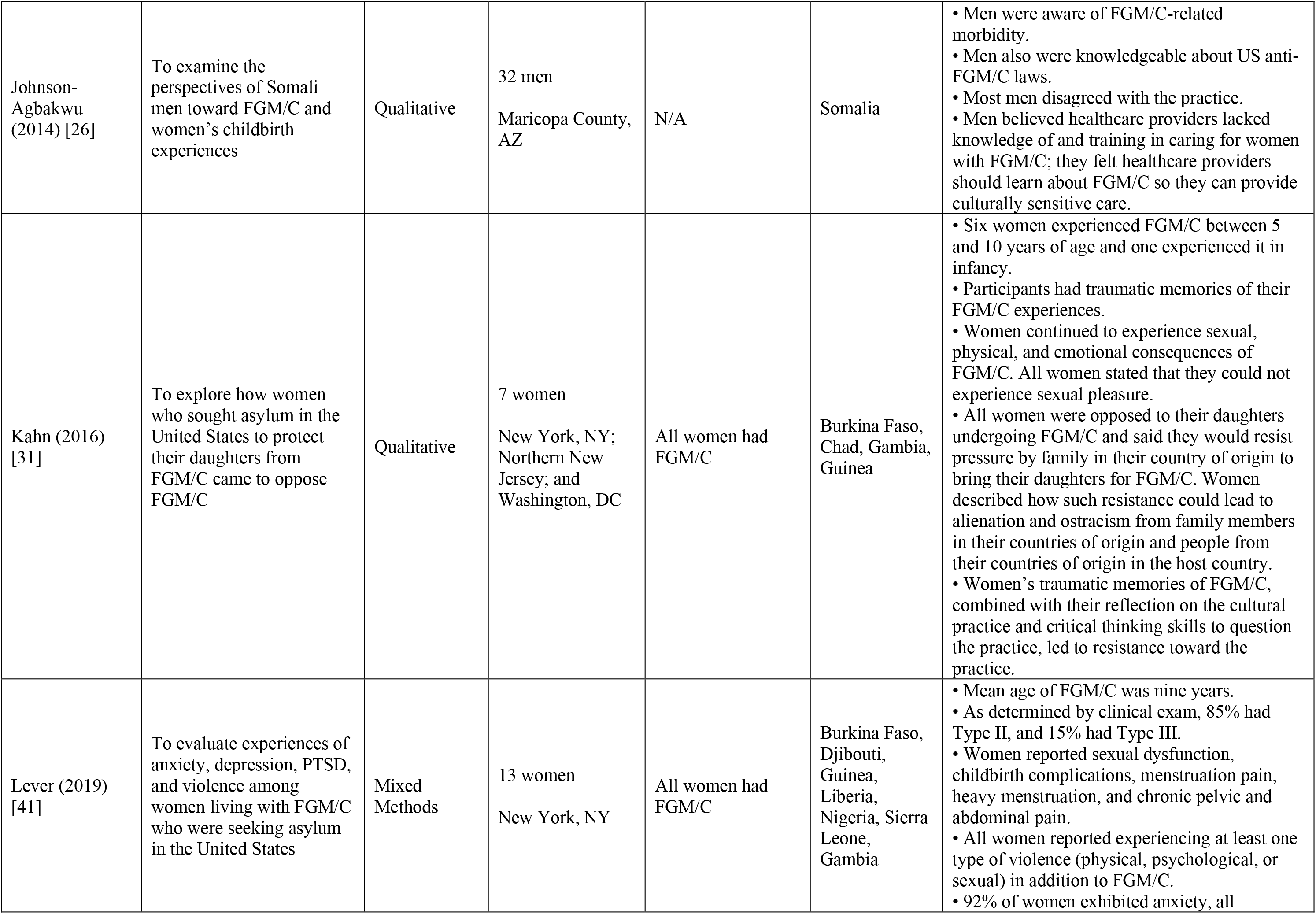
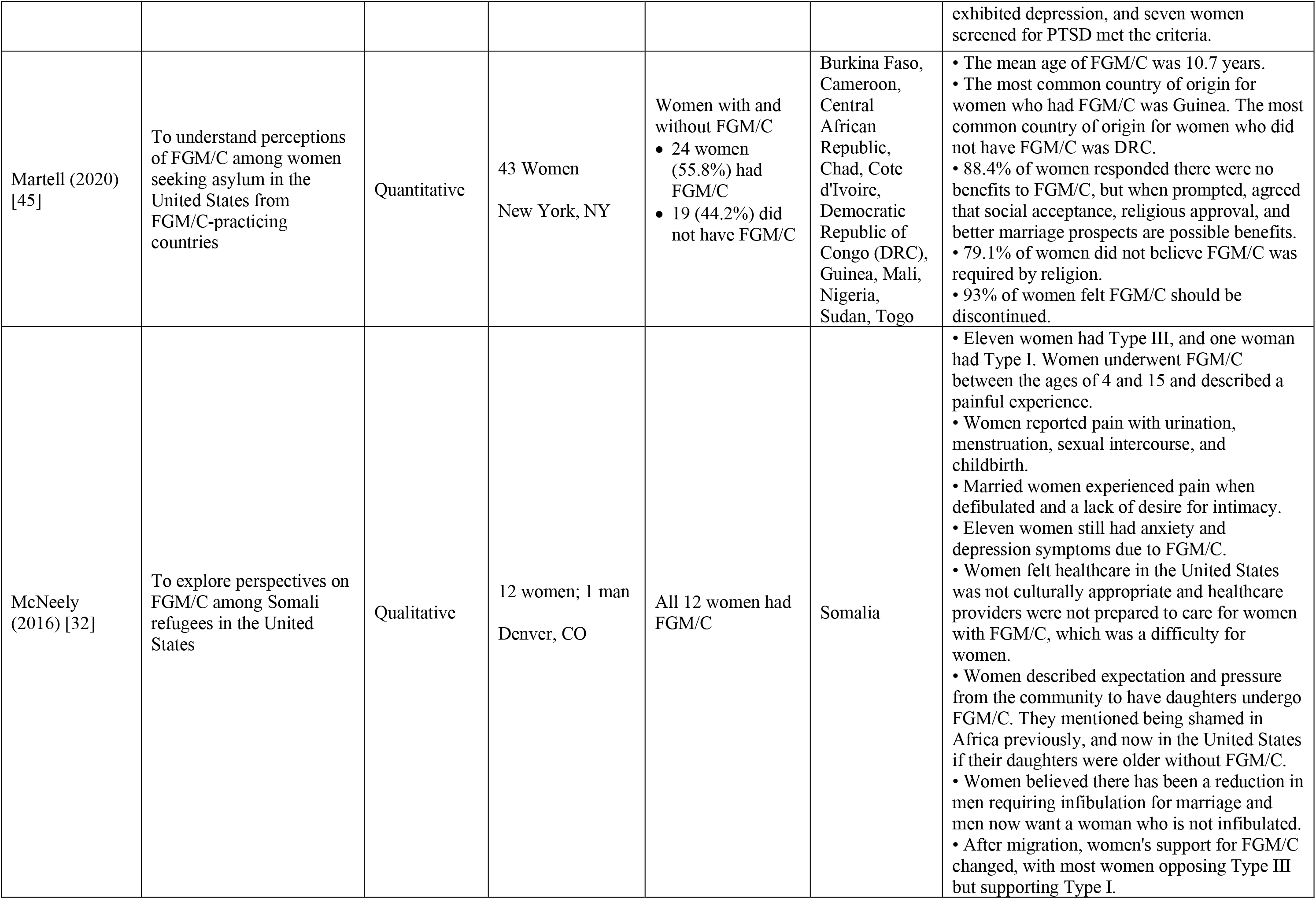
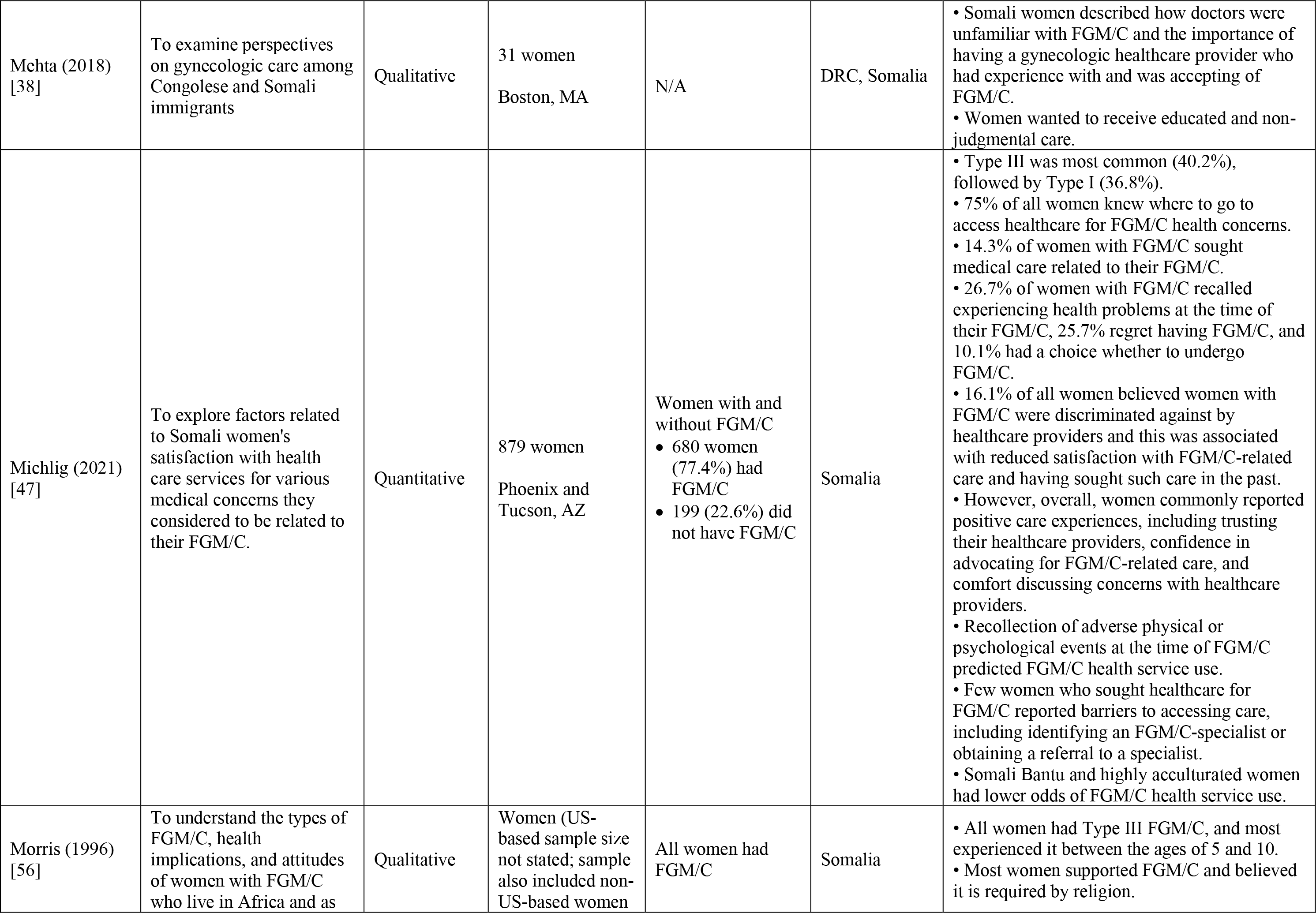
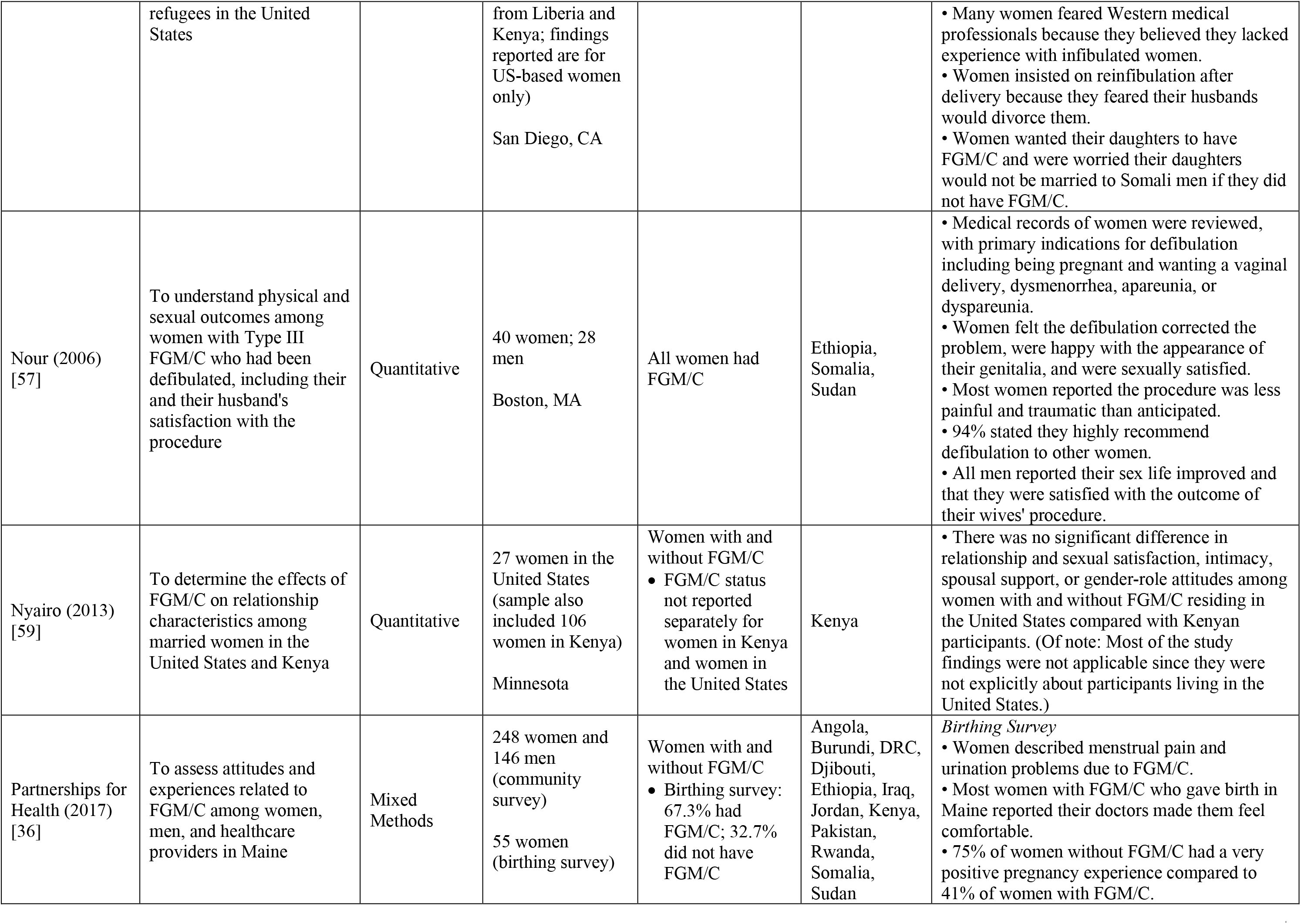
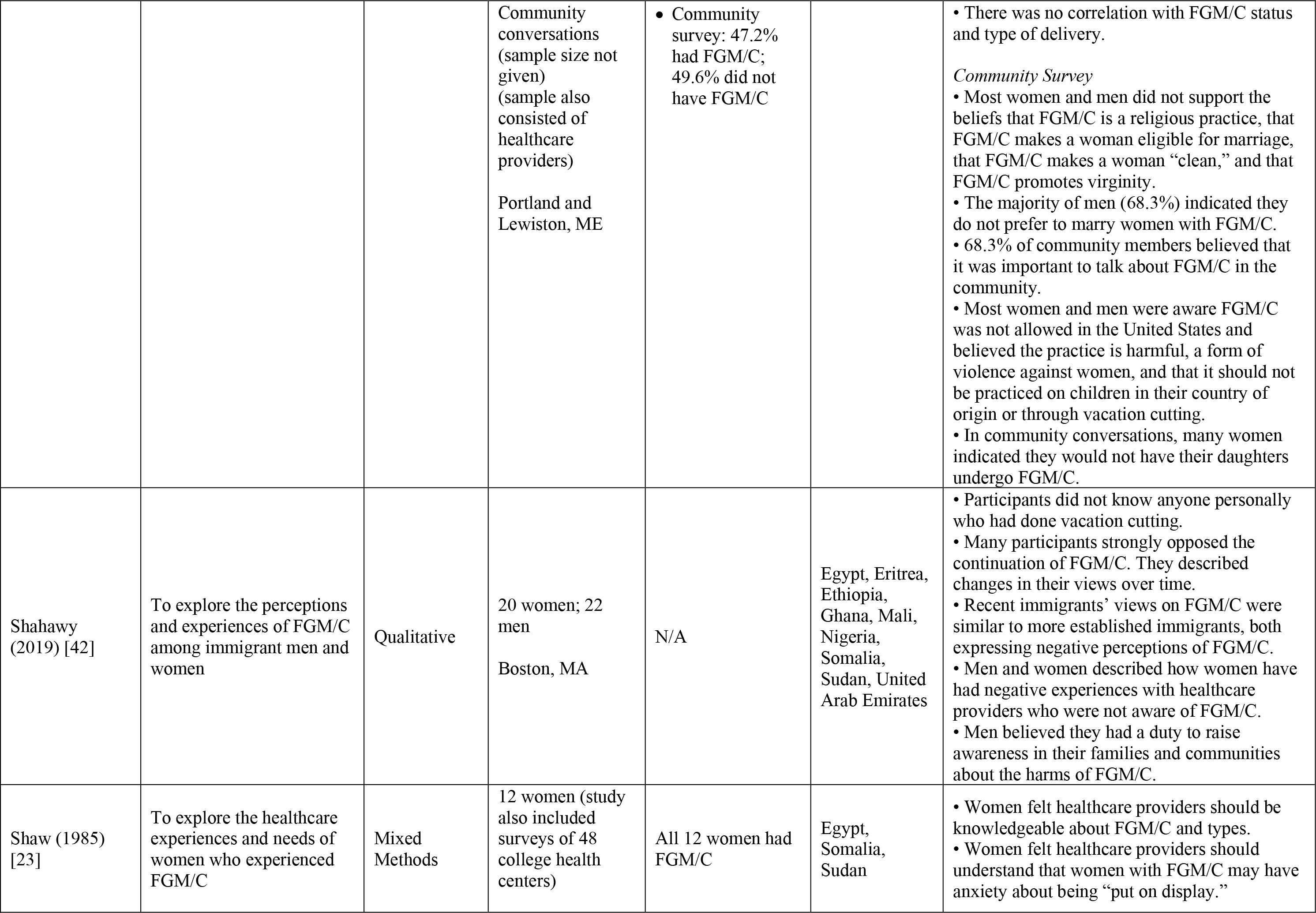
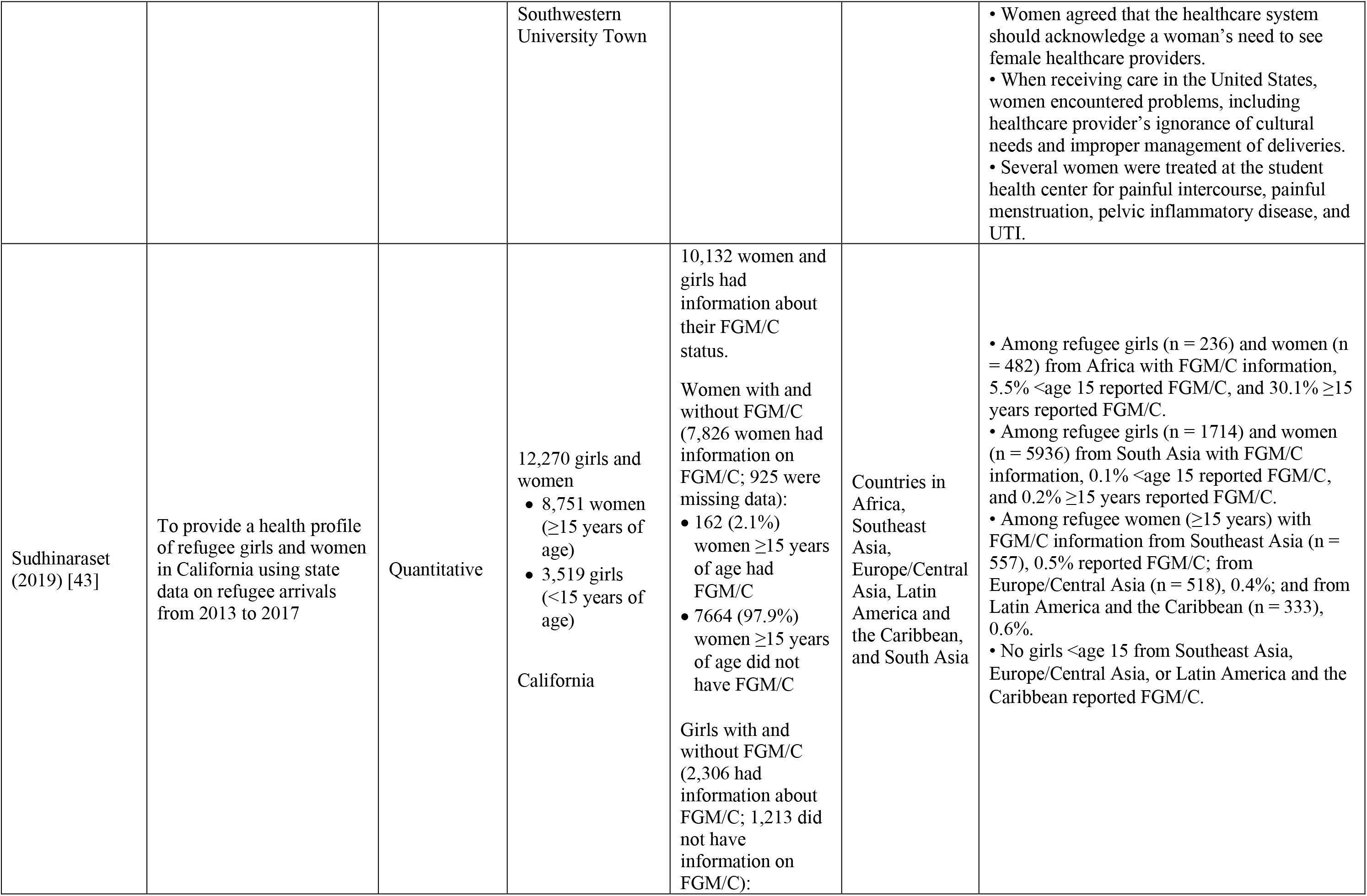
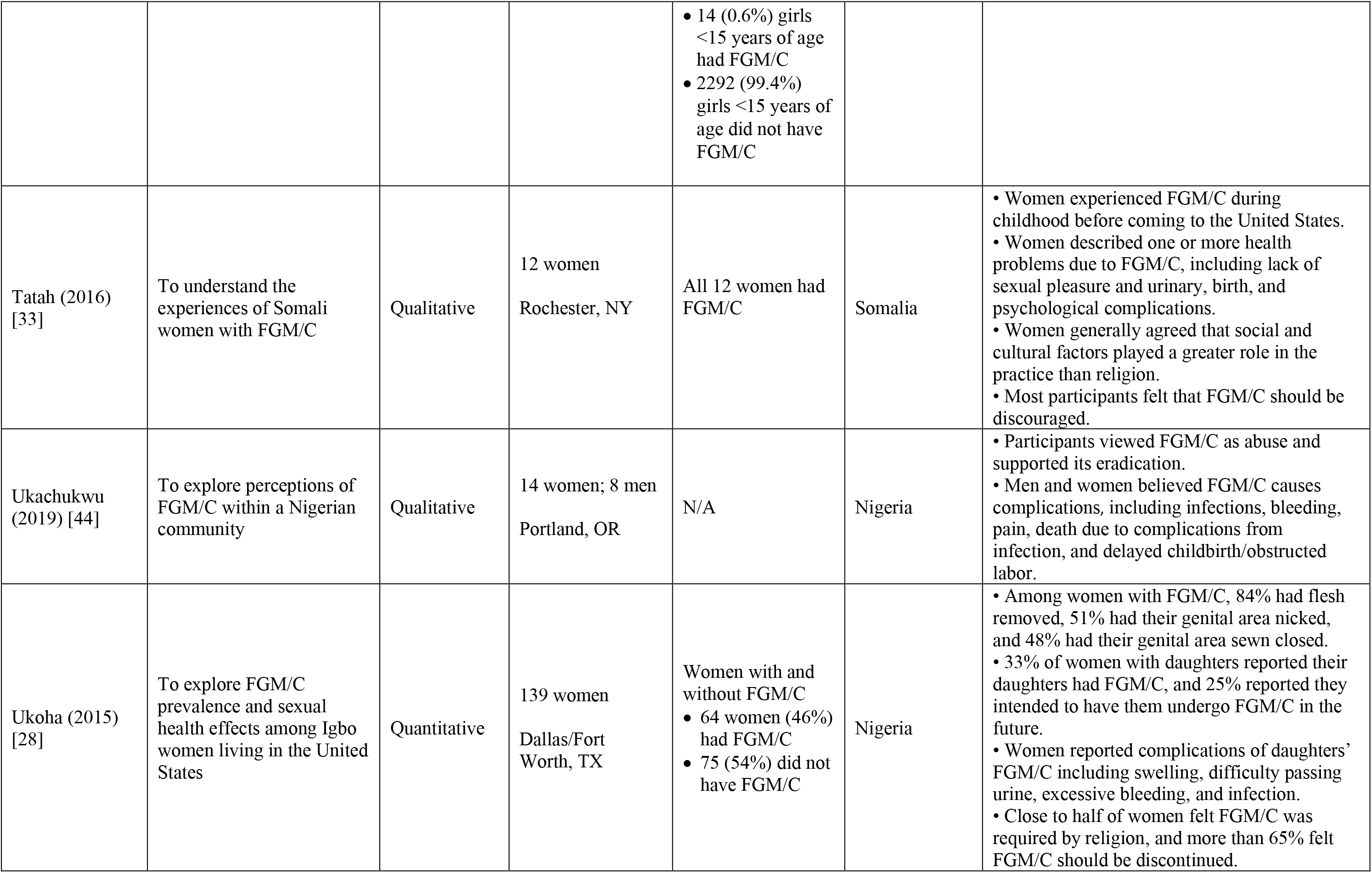
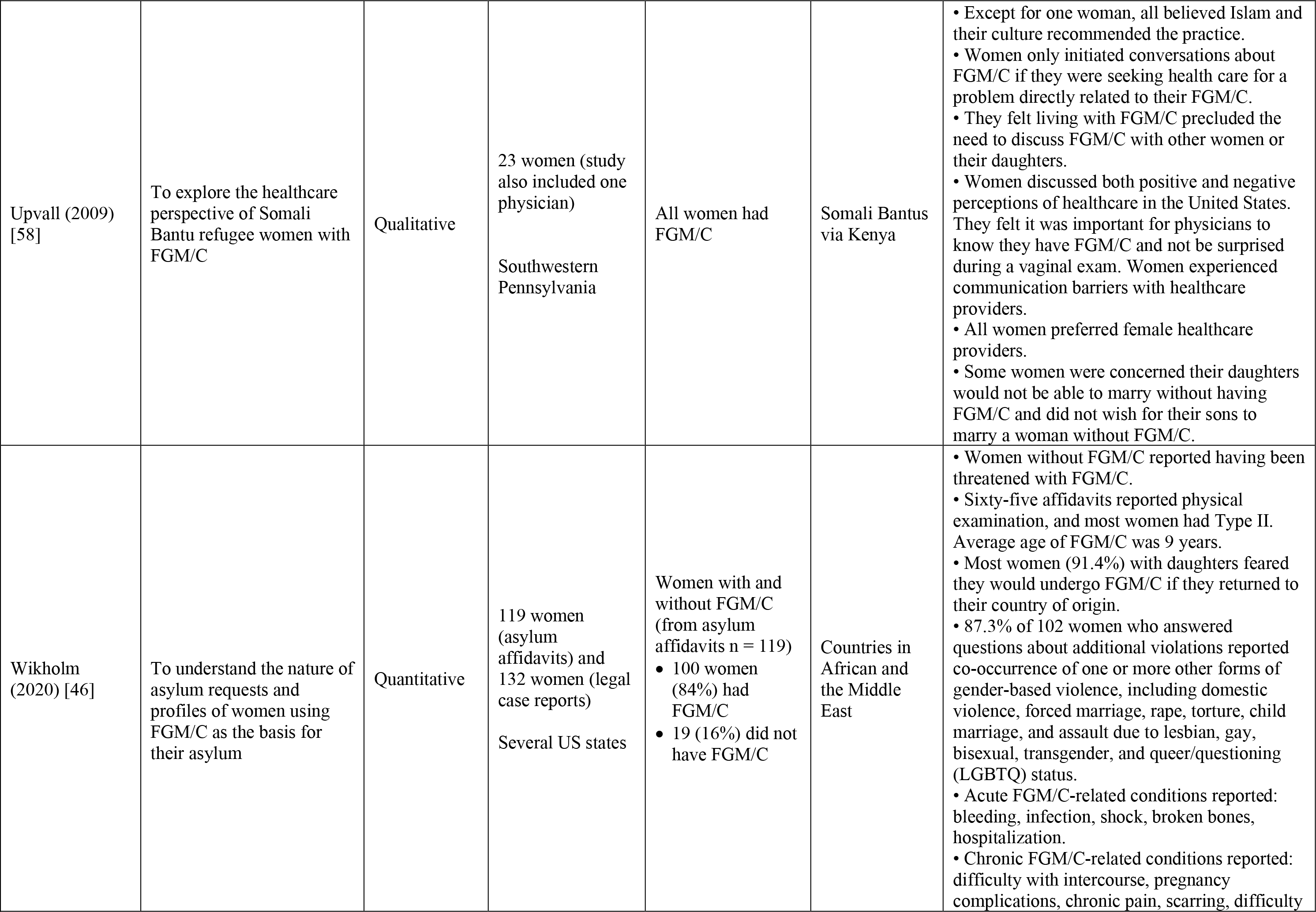
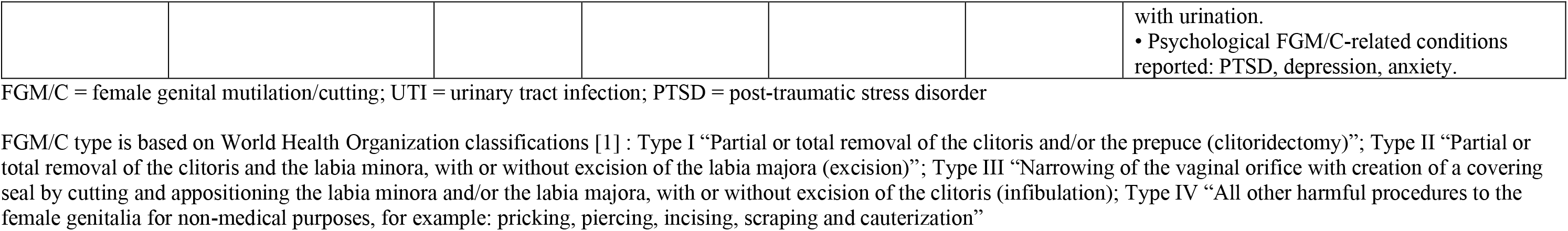
Included Studies.

| Lead Author (Year) | Aim(s) | Study Design | Study Population and Location | FGM/C Status of Study Population | Country(ies) of Origin | Main Findings Related to FGM/C |
| --- | --- | --- | --- | --- | --- | --- |
| Akinsulure-Smith (2014) [24] | To examine the FGM/C experience of West African immigrant women | Quantitative | 23 women<br>New York, NY | Women with and without FGM/C<br>• 7 women had FGM/C<br>• 16 did not have FGM/C | Liberia and Sierra Leone | • Muslim women had higher rates of FGM/C than Christian women, and FGM/C was more common among Sierra Leonean women compared with Liberian women.<br>• Psychological symptoms scores did not differ significantly between women with and without FGM/C; both groups experienced a similar number of traumatic life events. |
| Akinsulure-Smith (2017a) [34] | To explore knowledge and attitudes toward FGM/C among male West African immigrants | Quantitative | 36 men; 71 women<br>New York, NY | N/A | Gambia, Guinea, Mali, and Sierra Leone | • Men and women had similar levels of knowledge about FGM/C.<br>• Most men and women believed FGM/C should be stopped and did not intend to have their daughters undergo FGM/C.<br>• Most men had no preference for dating or marrying women with or without FGM/C. Men with a preference more commonly preferred women without FGM/C. |
| Akinsulure-Smith (2017b) [35] | To document the prevalence of FGM/C among refugee survivors of torture and trauma; and to compare characteristics, torture experiences, and mental health outcomes of women with and without FGM/C | Quantitative | 514 women<br>New York, NY | Women with and without FGM/C<br>• 133 women (25.9%) had FGM/C<br>• 381 (74.1%) did not have FGM/C | Multiple African countries, the majority West African | • A significantly greater proportion of women who experienced FGM/C were from West African countries, compared to other countries, and Muslim women compared to those who were Christian or other religions.<br>• Women's FGM/C status was confirmed during their gynecological exam once admitted to the clinic for survivors of torture.<br>• Women with and without FGM/C did not have significantly different Harvard Trauma Questionnaire scores.<br>• A larger proportion of women with FGM/C reported psychological and sexual torture than those without FGM/C. |
| Akinsulure-Smith (2018) [37] | To understand the experiences and attitudes of West African women immigrants who experienced FGM/C | Qualitative | 9 women<br>New York, NY | All women had FGM/C | Gambia, Guinea, and Mali | • Women described the primary rationale for FGM/C as being to reduce women's sexual pleasure to prevent premarital sex.<br>• Experience of FGM/C was traumatic, and themes that emerged included secrecy, surprise, and force.<br>• Women described short- and long-term physical consequences of FGM/C, including sexual arousal and childbirth difficulties.<br>• All women were against the practice.<br>• Women also discussed the persistent threat and fear of FGM/C without parent's consent, particularly when their daughters are in their home countries. They also described warnings from US doctors that if their daughters came back to the United States with FGM/C they would face legal consequences. One woman discussed her awareness of an instance of vacation cutting (transporting an individual from the United States to other countries for FGM/C).<br>• Women expressed frustrations that family members in home countries have difficulty understanding their refusal to perform FGM/C on girls in the United States.<br>• Experiences with healthcare providers varied. Some women hesitated to seek care because of fear of getting in trouble due to daughters' FGM/C or their undocumented status. |
| Ameresekere (2011) [52] | To explore perceptions of cesarean delivery and patient-provider communication related to FGM/C | Qualitative | 23 women<br>Boston, MA | All women had FGM/C | Somalia | • Women reported their healthcare providers in the United States did not discuss FGM/C. Fifteen women reported their physician did not mention FGM/C, 6 women said their physician acknowledged their FGM/C but did not discuss it in the context of their delivery, and 5 women said they wished their healthcare providers discussed their FGM/C. |
| Anuforo (2004) [51] | To gain an understanding of the meanings, beliefs, and practices related to FGM/C among Nigerians in the United States and Nigeria | Qualitative | 20 women and men (sample also included 30 women and men in Nigeria; results not broken out by sex)<br>Essex County, NJ | Women with and without FGM/C<br>• 14 US-based women had FGM/C<br>• Since results not broken out by sex, not clear how many women did not have FGM/C | Nigeria | • Fourteen US participants reported they experienced FGM/C, with FGM/C happening during infancy.<br>• Most US participants said they did not want FGM/C to continue. |
| Arbesman (1993) [48] | To determine whether the clinical implications of FGM/C were consistent with what was reported in the literature | Quantitative | 12 women<br>Buffalo, NY | Women with and without FGM/C<br>• 11 women had FGM/C<br>• 1 did not have FGM/C | Somalia | • Seven women with FGM/C experienced Type III. The mean age of FGM/C was 7.4 years.<br>• More than half of the women past the age of menarche reported pain during menstruation.<br>• Some women reported urination issues and pain during intercourse.<br>• No women reported difficulty getting pregnant.<br>• Half of women felt FGM/C was good.<br>• Seven women said they support milder forms of FGM/C for their daughters. |
| Banke-Thomas (2019) [39] | To assess factors that influence Somali women's access to maternal and reproductive health | Quantitative | 427 women<br>Franklin County, OH | Women with and without FGM/C<br>• 336 women (82.2%) had FGM/C<br>• 73 (17.8%) did not have FGM/C | Somalia | • More than half (58.1%) of women with FGM/C reported Type III.<br>• Women with FGM/C were less willing to seek maternal and reproductive health care.<br>• FGM/C status was not associated with gaining entry to the health system, seeing a primary provider, or seeing a specialist.<br>• Women with more severe forms of FGM/C were less willing to seek care and had more difficulty in gaining entry to the health system and in accessing a primary provider compared to women with Type I. |
| Beine (1995) [53] | To understand Somali women's cultural beliefs and behaviors during pregnancy and their attitudes toward prenatal care experiences | Qualitative | 14 women<br>San Diego, CA | Women with and without FGM/C<br>• Exact number not known because some women were uncomfortable disclosing | Somalia | • Women described feeling humiliated by healthcare providers' reactions to their FGM/C.<br>• One woman indicated she intended to go back to Somalia for her daughter's FGM/C. Others were hesitant to express their intentions.<br>• Women were generally satisfied with the quality of prenatal care they received. When seeking prenatal care, women wanted a female healthcare provider who was informed about FGM/C. |
| Brown (2010) [49] | To explore reasons for Somali women's resistance to prenatal and obstetrical interventions in the United States | Qualitative | 34 women<br>Rochester, NY | N/A | Somalia | • Some women supported the continuance of FGM/C while others thought it should stop.<br>• Women had diverse opinions about whether FGM/C causes health problems.<br>• Women described health problems that occur immediately after FGM/C and problems later in life, including urination problems, menstruation difficulties, painful intercourse, and needing "cutting" during delivery. |
|  |  |  |  |  |  | • No women associated FGM/C with the need for a cesarean section. |
| Chu (2016) [29] | To examine the prevalence of FGM/C; reproductive sexual function, and mental health outcomes associated with FGM/C; and attitudes toward FGM/C among West African immigrant women | Quantitative | 68 women<br>New York, NY | Women with and without FGM/C<br>• 46 women (68%) had FGM/C<br>• 22 (32%) did not have FGM/C | Gambia, Guinea, Mali, Sierra Leone | • The percentage of women with FGM/C ranged from 45% among Sierra Leonean women to 92% among Malian women.<br>• Most common age of FGM/C was between 6 and 10 years.<br>• 8.9% reported no flesh removed, 91.9% reported flesh removed, and 40% reported their genital area was sewn closed.<br>• 86.2% of women felt FGM/C should be stopped.<br>• 94.8% of women did not intend to have their daughters undergo FGM/C.<br>• Women with FGM/C had significantly more live births and reported more vaginal pain and decreased sexual arousal compared with those without FGM/C.<br>• There were no other differences between women with and without FGM/C in other gynecological and obstetric, sexual, and mental health outcomes measured. |
| Connor (2016) [30] | To investigate the sexual values, attitudes, and behaviors of female Somali refugees | Mixed-Methods | 30 women<br>Minnesota | Women with and without FGM/C<br>• 28 women had FGM/C<br>• 2 did not have FGM/C | Somalia | • 63% of women with FGM/C had Type II or III, and 37% had Type I or IV.<br>• Half of the women stated that FGM/C negatively affected their sexual relationship.<br>• Women with Type II or III were more likely to report having pain with vaginal intercourse than those who had Type I or IV.<br>• 85% of women indicated they did not or would not have their daughters undergo FGM/C and reported negative attitudes about FGM/C. |
| Dahlen (2011) [60] | To understand women's experiences of growing up in Ethiopia, being an immigrant mother in the United States, and making meaning of her experiences, particularly as it relates to FGM/C | Qualitative | 9 women<br>Not specified | Women with and without FGM/C<br>• 7 women had FGM/C<br>• 2 did not have FGM/C | Ethiopia | • Women expressed that they had feelings of physical and emotional pain, including helplessness and anger, during their FGM/C.<br>• Women described a lack of sexual pleasure because of FGM/C.<br>• Women opposed FGM/C and did not want their daughters to experience it.<br>• Women had varied views about whether to tell their children about FGM/C.<br>• Women described abandoning FGM/C after immigration to the United States.<br>• Most women were aware FGM/C was illegal in the United States; some were grateful the practice was illegal, so that they can protect their daughters from the practice. |
| Deason (2013) [54] | To understand African women's perceptions of FGM/C and FGM/C laws | Qualitative | 9 women<br>Maryland, Minnesota, New York, North Carolina, and Ohio | N/A | Egypt, Kenya, Somalia | • Women reported a variety of reasons why their communities practiced FGM/C, including preventing premarital sex, marriageability, and reducing sexual urges.<br>• All women expressed they were aware of the health problems associated with FGM/C, and they were against the practice.<br>• Several women reported adverse health outcomes because of FGM/C.<br>• All women were generally aware of FGM/C laws in the United States. None of the women felt anti-FGM/C legislation was effective. |
| Eyega (1997) [50] | To determine the extent of FGM/C in African immigrant communities in New York City, their social service needs, and the training and information needs of their healthcare providers | Quantitative | 20 women (study also included healthcare providers)<br>New York, NY | Women with and without FGM/C<br>• 18 women had FGM/C<br>• 2 did not have FGM/C | Egypt, Ethiopia, Gambia, Guinea, Mali, Nigeria, Sierra Leone, Sudan | • Eight remembered their age of FGM/C, and this ranged from 3 to 18 years.<br>• No women reported their daughters had FGM/C.<br>• Ten women were aware of problems associated with FGM/C, primarily mentioning excessive bleeding and difficulty during intercourse.<br>• Four women reported recurrent gynecological issues during the last 2 years; they all had Type III, and three reported these problems affected their marriage.<br>• Fourteen women were very or somewhat satisfied with the health services they received, compared to five who reported they were somewhat or very dissatisfied. |
| Fawcett (2014) [25] | To explore patterns of cultural agreement and variation in knowledge in childbearing between Somali refugees and healthcare providers and their perceptions of FGM/C | Mixed-Methods | 73 women (study also included healthcare providers)<br>Phoenix, AZ | Women with and without FGM/C<br>• 67 women (91.7%) had FGM/C<br>• 4 (5.5%) did not have FGM/C | Somalia | • 42 women with FGM/C had Type III, and 24 had Type I or II.<br>• Among the different Somali populations, FGM/C is central to their traditional beliefs and practices, although there are differences in managing FGM/C during childbirth.<br>• Most women did not believe that FGM/C is associated with increased risks of adverse birth outcomes. |
|  |  |  |  |  |  | • Most women were critical of FGM/C, especially Type III. Some women viewed less severe forms as acceptable.<br>• A theme that emerged among women was fear and distrust of medical care in the United States. |
| Fox (2020) [9] | To examine if exposure to victimization (home or property looted or burned, abandoned or thrown out by family, abducted, experienced sexual violence, attacked with a weapon, or witnessed a murder) is related to health problems, healthcare access and barriers, and unmet healthcare needs among Somali women and adolescent girls, after accounting for the effects of FGM/C | Quantitative | 879 women<br>Phoenix and Tucson, Arizona | Women with and without FGM/C<br>• 687 women (78%) had FGM/C<br>• 192 (21%) did not have FGM/C | Somalia | • Among non-single women, FGM/C was significantly associated with problems with sexual intercourse and pregnancy and with having had a Pap test.<br>• Among all women, FGM/C was associated with having a designated place to receive healthcare, feelings of depression or flashbacks/nightmares about a traumatic event, and gynecological issues. Among women with FGM/C, the predicted probability of having a designated healthcare place was lower among those exposed to victimization compared to those who were not.<br>• Among women with FGM/C, those who were exposed to victimization had a higher predicted probability of pregnancy-related problems, gynecological issues, and unmet healthcare needs compared to women with FGM/C who were not exposed to victimization. |
| Gali (1997) [61] | To explore the relationship between psychological and medical concerns women with FGM/C encounter and barriers to reproductive healthcare in the United States | Qualitative | 50 women<br>Bay Area, CA | Women with and without FGM/C<br>• 42% of women directly indicated they had FGM/C<br>• Exact number not known because some women were reluctant to discuss | Egypt, Eritrea, Ethiopia, Sudan | • Most women (80%) did not intend to have their daughters undergo FGM/C because they felt it was not mandatory for their religion. Those who did reported they would do so to follow other family members' desires and so their daughters can have a “good” future marriage.<br>• All women reported at least one negative experience in the US healthcare system related to culturally sensitive care and FGM/C.<br>• Some women said they were uncomfortable with how they were treated and preferred healthcare providers familiar with FGM/C. |
| Geynisman-Tan (2019) [40] | To describe symptom prevalence and quality of life impact from lower urinary tract symptoms among women with FGM/C in the United States | Quantitative | 30 women<br>Boston, MA; Chicago, IL; New York, NY; Minneapolis, MN; and San Francisco, CA | All women had FGM/C | Not specified | • 40% of women reported Type I, 23% Type II, 23% Type III, and 13% were unsure.<br>• Women reported experiencing FGM/C between the ages of one week and 16 years.<br>• A history of UTIs was common. 46% reported at least one UTI since FGM/C, 26% in the last year, and 10% reported more than three UTIs in the last year.<br>• 73% were positive for experiencing at least one lower urinary tract symptom sometimes, most of the time, or all the time. Urgency was the most commonly reported symptom (60%), followed by urgency urinary incontinence (53%).<br>• 63% reported that urinary symptoms had a “moderate” or “quite a bit of” impact on their activities, relationships, or feelings. |
| Horowitz (1997) [55] | To understand the conflicts that occur when healthcare providers care for patients whose beliefs and practices differ from their own | Qualitative | 45 women; 5 men (study also included healthcare providers)<br>US location(s) not specified | All women had FGM/C | Eritrea, Ethiopia, Somalia | • All Somali women had infibulation (Type III); Eritrean and Ethiopian women had FGM/C types ranging from Type I to Type III.<br>• Some men opposed FGM/C. However, some women did not trust men’s opinions that they did not care if women had FGM/C.<br>• Women with FGM/C were offended when their healthcare providers invited colleagues to view the woman’s genitalia.<br>• Some women feared that girls without FGM/C would not be culturally accepted, while other women were eager to stop the practice.<br>• Some women recalled their FGM/C as being traumatic and painful; other women did not remember or only recalled a celebration. |
| Jacoby (2015) [27] | To assess Somali women’s health literacy and perinatal experiences | Mixed Methods | 23 women<br>Lewiston, ME | Not clear | Somalia | • Women with FGM/C reported problems with their FGM/C related to childbirth, sex, pain, UTIs, and menstruation.<br>• Most women did not want their daughters to undergo FGM/C because of the impacts it can have on a woman’s body. |
| Johnson-Agbakwu (2014) [26] | To examine the perspectives of Somali men toward FGM/C and women's childbirth experiences | Qualitative | 32 men<br>Maricopa County, AZ | N/A | Somalia | • Men were aware of FGM/C-related morbidity.<br>• Men also were knowledgeable about US anti-FGM/C laws.<br>• Most men disagreed with the practice.<br>• Men believed healthcare providers lacked knowledge of and training in caring for women with FGM/C; they felt healthcare providers should learn about FGM/C so they can provide culturally sensitive care. |
| Kahn (2016) [31] | To explore how women who sought asylum in the United States to protect their daughters from FGM/C came to oppose FGM/C | Qualitative | 7 women<br>New York, NY; Northern New Jersey; and Washington, DC | All women had FGM/C | Burkina Faso, Chad, Gambia, Guinea | • Six women experienced FGM/C between 5 and 10 years of age and one experienced it in infancy.<br>• Participants had traumatic memories of their FGM/C experiences.<br>• Women continued to experience sexual, physical, and emotional consequences of FGM/C. All women stated that they could not experience sexual pleasure.<br>• All women were opposed to their daughters undergoing FGM/C and said they would resist pressure by family in their country of origin to bring their daughters for FGM/C. Women described how such resistance could lead to alienation and ostracism from family members in their countries of origin and people from their countries of origin in the host country.<br>• Women's traumatic memories of FGM/C, combined with their reflection on the cultural practice and critical thinking skills to question the practice, led to resistance toward the practice. |
| Lever (2019) [41] | To evaluate experiences of anxiety, depression, PTSD, and violence among women living with FGM/C who were seeking asylum in the United States | Mixed Methods | 13 women<br>New York, NY | All women had FGM/C | Burkina Faso, Djibouti, Guinea, Liberia, Nigeria, Sierra Leone, Gambia | • Mean age of FGM/C was nine years.<br>• As determined by clinical exam, 85% had Type II, and 15% had Type III.<br>• Women reported sexual dysfunction, childbirth complications, menstruation pain, heavy menstruation, and chronic pelvic and abdominal pain.<br>• All women reported experiencing at least one type of violence (physical, psychological, or sexual) in addition to FGM/C.<br>• 92% of women exhibited anxiety, all exhibited depression, and seven women screened for PTSD met the criteria. |
| Martell (2020) [45] | To understand perceptions of FGM/C among women seeking asylum in the United States from FGM/C-practicing countries | Quantitative | 43 Women<br>New York, NY | Women with and without FGM/C<br>• 24 women (55.8%) had FGM/C<br>• 19 (44.2%) did not have FGM/C | Burkina Faso, Cameroon, Central African Republic, Chad, Cote d'Ivoire, Democratic Republic of Congo (DRC), Guinea, Mali, Nigeria, Sudan, Togo | • The mean age of FGM/C was 10.7 years.<br>• The most common country of origin for women who had FGM/C was Guinea. The most common country of origin for women who did not have FGM/C was DRC.<br>• 88.4% of women responded there were no benefits to FGM/C, but when prompted, agreed that social acceptance, religious approval, and better marriage prospects are possible benefits.<br>• 79.1% of women did not believe FGM/C was required by religion.<br>• 93% of women felt FGM/C should be discontinued. |
| McNeely (2016) [32] | To explore perspectives on FGM/C among Somali refugees in the United States | Qualitative | 12 women; 1 man<br>Denver, CO | All 12 women had FGM/C | Somalia | • Eleven women had Type III, and one woman had Type I. Women underwent FGM/C between the ages of 4 and 15 and described a painful experience.<br>• Women reported pain with urination, menstruation, sexual intercourse, and childbirth.<br>• Married women experienced pain when defibulated and a lack of desire for intimacy.<br>• Eleven women still had anxiety and depression symptoms due to FGM/C.<br>• Women felt healthcare in the United States was not culturally appropriate and healthcare providers were not prepared to care for women with FGM/C, which was a difficulty for women.<br>• Women described expectation and pressure from the community to have daughters undergo FGM/C. They mentioned being shamed in Africa previously, and now in the United States if their daughters were older without FGM/C.<br>• Women believed there has been a reduction in men requiring infibulation for marriage and men now want a woman who is not infibulated.<br>• After migration, women's support for FGM/C changed, with most women opposing Type III but supporting Type I. |
| Mehta (2018)<br>[38] | To examine perspectives on gynecologic care among Congolese and Somali immigrants | Qualitative | 31 women<br>Boston, MA | N/A | DRC, Somalia | • Somali women described how doctors were unfamiliar with FGM/C and the importance of having a gynecologic healthcare provider who had experience with and was accepting of FGM/C.<br>• Women wanted to receive educated and non-judgmental care. |
| Michlig (2021)<br>[47] | To explore factors related to Somali women's satisfaction with health care services for various medical concerns they considered to be related to their FGM/C. | Quantitative | 879 women<br>Phoenix and Tucson, AZ | Women with and without FGM/C<br>• 680 women (77.4%) had FGM/C<br>• 199 (22.6%) did not have FGM/C | Somalia | • Type III was most common (40.2%), followed by Type I (36.8%).<br>• 75% of all women knew where to go to access healthcare for FGM/C health concerns.<br>• 14.3% of women with FGM/C sought medical care related to their FGM/C.<br>• 26.7% of women with FGM/C recalled experiencing health problems at the time of their FGM/C, 25.7% regret having FGM/C, and 10.1% had a choice whether to undergo FGM/C.<br>• 16.1% of all women believed women with FGM/C were discriminated against by healthcare providers and this was associated with reduced satisfaction with FGM/C-related care and having sought such care in the past.<br>• However, overall, women commonly reported positive care experiences, including trusting their healthcare providers, confidence in advocating for FGM/C-related care, and comfort discussing concerns with healthcare providers.<br>• Recollection of adverse physical or psychological events at the time of FGM/C predicted FGM/C health service use.<br>• Few women who sought healthcare for FGM/C reported barriers to accessing care, including identifying an FGM/C-specialist or obtaining a referral to a specialist.<br>• Somali Bantu and highly acculturated women had lower odds of FGM/C health service use. |
| Morris (1996)<br>[56] | To understand the types of FGM/C, health implications, and attitudes of women with FGM/C who live in Africa and as | Qualitative | Women (US-based sample size not stated; sample also included non-US-based women | All women had FGM/C | Somalia | • All women had Type III FGM/C, and most experienced it between the ages of 5 and 10.<br>• Most women supported FGM/C and believed it is required by religion. |
|  | refugees in the United States |  | from Liberia and Kenya; findings reported are for US-based women only)<br>San Diego, CA |  |  | • Many women feared Western medical professionals because they believed they lacked experience with infibulated women.<br>• Women insisted on reinfibulation after delivery because they feared their husbands would divorce them.<br>• Women wanted their daughters to have FGM/C and were worried their daughters would not be married to Somali men if they did not have FGM/C. |
| Nour (2006) [57] | To understand physical and sexual outcomes among women with Type III FGM/C who had been defibulated, including their and their husband's satisfaction with the procedure | Quantitative | 40 women; 28 men<br>Boston, MA | All women had FGM/C | Ethiopia, Somalia, Sudan | • Medical records of women were reviewed, with primary indications for defibulation including being pregnant and wanting a vaginal delivery, dysmenorrhea, apareunia, or dyspareunia.<br>• Women felt the defibulation corrected the problem, were happy with the appearance of their genitalia, and were sexually satisfied.<br>• Most women reported the procedure was less painful and traumatic than anticipated.<br>• 94% stated they highly recommend defibulation to other women.<br>• All men reported their sex life improved and that they were satisfied with the outcome of their wives' procedure. |
| Nyairo (2013) [59] | To determine the effects of FGM/C on relationship characteristics among married women in the United States and Kenya | Quantitative | 27 women in the United States (sample also included 106 women in Kenya)<br>Minnesota | Women with and without FGM/C<br>• FGM/C status not reported separately for women in Kenya and women in the United States | Kenya | • There was no significant difference in relationship and sexual satisfaction, intimacy, spousal support, or gender-role attitudes among women with and without FGM/C residing in the United States compared with Kenyan participants. (Of note: Most of the study findings were not applicable since they were not explicitly about participants living in the United States.) |
| Partnerships for Health (2017) [36] | To assess attitudes and experiences related to FGM/C among women, men, and healthcare providers in Maine | Mixed Methods | 248 women and 146 men (community survey)<br>55 women (birthing survey) | Women with and without FGM/C<br>• Birthing survey: 67.3% had FGM/C; 32.7% did not have FGM/C | Angola, Burundi, DRC, Djibouti, Ethiopia, Iraq, Jordan, Kenya, Pakistan, Rwanda, Somalia, Sudan | *Birthing Survey*<br>• Women described menstrual pain and urination problems due to FGM/C.<br>• Most women with FGM/C who gave birth in Maine reported their doctors made them feel comfortable.<br>• 75% of women without FGM/C had a very positive pregnancy experience compared to 41% of women with FGM/C. |
|  |  |  | Community conversations (sample size not given) (sample also consisted of healthcare providers)<br>Portland and Lewiston, ME | Community survey: 47.2% had FGM/C; 49.6% did not have FGM/C |  | There was no correlation with FGM/C status and type of delivery.<br>*Community Survey*<br>Most women and men did not support the beliefs that FGM/C is a religious practice, that FGM/C makes a woman eligible for marriage, that FGM/C makes a woman “clean,” and that FGM/C promotes virginity.<br>The majority of men (68.3%) indicated they do not prefer to marry women with FGM/C.<br>68.3% of community members believed that it was important to talk about FGM/C in the community.<br>Most women and men were aware FGM/C was not allowed in the United States and believed the practice is harmful, a form of violence against women, and that it should not be practiced on children in their country of origin or through vacation cutting.<br>In community conversations, many women indicated they would not have their daughters undergo FGM/C. |
| Shahawy (2019) [42] | To explore the perceptions and experiences of FGM/C among immigrant men and women | Qualitative | 20 women; 22 men<br>Boston, MA | N/A | Egypt, Eritrea, Ethiopia, Ghana, Mali, Nigeria, Somalia, Sudan, United Arab Emirates | Participants did not know anyone personally who had done vacation cutting.<br>Many participants strongly opposed the continuation of FGM/C. They described changes in their views over time.<br>Recent immigrants’ views on FGM/C were similar to more established immigrants, both expressing negative perceptions of FGM/C.<br>Men and women described how women have had negative experiences with healthcare providers who were not aware of FGM/C.<br>Men believed they had a duty to raise awareness in their families and communities about the harms of FGM/C. |
| Shaw (1985) [23] | To explore the healthcare experiences and needs of women who experienced FGM/C | Mixed Methods | 12 women (study also included surveys of 48 college health centers) | All 12 women had FGM/C | Egypt, Somalia, Sudan | Women felt healthcare providers should be knowledgeable about FGM/C and types.<br>Women felt healthcare providers should understand that women with FGM/C may have anxiety about being “put on display.” |
|  |  |  | Southwestern University Town |  |  | • Women agreed that the healthcare system should acknowledge a woman's need to see female healthcare providers.<br>• When receiving care in the United States, women encountered problems, including healthcare provider's ignorance of cultural needs and improper management of deliveries.<br>• Several women were treated at the student health center for painful intercourse, painful menstruation, pelvic inflammatory disease, and UTI. |
| Sudhinaraset (2019) [43] | To provide a health profile of refugee girls and women in California using state data on refugee arrivals from 2013 to 2017 | Quantitative | 12,270 girls and women<br>• 8,751 women (≥15 years of age)<br>• 3,519 girls (<15 years of age)<br>California | 10,132 women and girls had information about their FGM/C status.<br>Women with and without FGM/C (7,826 women had information on FGM/C; 925 were missing data):<br>• 162 (2.1%) women ≥15 years of age had FGM/C<br>• 7664 (97.9%) women ≥15 years of age did not have FGM/C<br>Girls with and without FGM/C (2,306 had information about FGM/C; 1,213 did not have information on FGM/C): | Countries in Africa, Southeast Asia, Europe/Central Asia, Latin America and the Caribbean, and South Asia | • Among refugee girls (n = 236) and women (n = 482) from Africa with FGM/C information, 5.5% <age 15 reported FGM/C, and 30.1% ≥15 years reported FGM/C.<br>• Among refugee girls (n = 1714) and women (n = 5936) from South Asia with FGM/C information, 0.1% <age 15 reported FGM/C, and 0.2% ≥15 years reported FGM/C.<br>• Among refugee women (≥15 years) with FGM/C information from Southeast Asia (n = 557), 0.5% reported FGM/C; from Europe/Central Asia (n = 518), 0.4%; and from Latin America and the Caribbean (n = 333), 0.6%.<br>• No girls <age 15 from Southeast Asia, Europe/Central Asia, or Latin America and the Caribbean reported FGM/C. |
|  |  |  |  | • 14 (0.6%) girls <15 years of age had FGM/C<br>• 2292 (99.4%) girls <15 years of age did not have FGM/C |  |  |
| Tatah (2016) [33] | To understand the experiences of Somali women with FGM/C | Qualitative | 12 women<br>Rochester, NY | All 12 women had FGM/C | Somalia | • Women experienced FGM/C during childhood before coming to the United States.<br>• Women described one or more health problems due to FGM/C, including lack of sexual pleasure and urinary, birth, and psychological complications.<br>• Women generally agreed that social and cultural factors played a greater role in the practice than religion.<br>• Most participants felt that FGM/C should be discouraged. |
| Ukachukwu (2019) [44] | To explore perceptions of FGM/C within a Nigerian community | Qualitative | 14 women; 8 men<br>Portland, OR | N/A | Nigeria | • Participants viewed FGM/C as abuse and supported its eradication.<br>• Men and women believed FGM/C causes complications, including infections, bleeding, pain, death due to complications from infection, and delayed childbirth/obstructed labor. |
| Ukoha (2015) [28] | To explore FGM/C prevalence and sexual health effects among Igbo women living in the United States | Quantitative | 139 women<br>Dallas/Fort Worth, TX | Women with and without FGM/C<br>• 64 women (46%) had FGM/C<br>• 75 (54%) did not have FGM/C | Nigeria | • Among women with FGM/C, 84% had flesh removed, 51% had their genital area nicked, and 48% had their genital area sewn closed.<br>• 33% of women with daughters reported their daughters had FGM/C, and 25% reported they intended to have them undergo FGM/C in the future.<br>• Women reported complications of daughters' FGM/C including swelling, difficulty passing urine, excessive bleeding, and infection.<br>• Close to half of women felt FGM/C was required by religion, and more than 65% felt FGM/C should be discontinued. |
| Upvall (2009) [58] | To explore the healthcare perspective of Somali Bantu refugee women with FGM/C | Qualitative | 23 women (study also included one physician)<br>Southwestern Pennsylvania | All women had FGM/C | Somali Bantus via Kenya | • Except for one woman, all believed Islam and their culture recommended the practice.<br>• Women only initiated conversations about FGM/C if they were seeking health care for a problem directly related to their FGM/C.<br>• They felt living with FGM/C precluded the need to discuss FGM/C with other women or their daughters.<br>• Women discussed both positive and negative perceptions of healthcare in the United States. They felt it was important for physicians to know they have FGM/C and not be surprised during a vaginal exam. Women experienced communication barriers with healthcare providers.<br>• All women preferred female healthcare providers.<br>• Some women were concerned their daughters would not be able to marry without having FGM/C and did not wish for their sons to marry a woman without FGM/C. |
| Wikholm (2020) [46] | To understand the nature of asylum requests and profiles of women using FGM/C as the basis for their asylum | Quantitative | 119 women (asylum affidavits) and 132 women (legal case reports)<br>Several US states | Women with and without FGM/C (from asylum affidavits n = 119)<br>• 100 women (84%) had FGM/C<br>• 19 (16%) did not have FGM/C | Countries in African and the Middle East | • Women without FGM/C reported having been threatened with FGM/C.<br>• Sixty-five affidavits reported physical examination, and most women had Type II. Average age of FGM/C was 9 years.<br>• Most women (91.4%) with daughters feared they would undergo FGM/C if they returned to their country of origin.<br>• 87.3% of 102 women who answered questions about additional violations reported co-occurrence of one or more other forms of gender-based violence, including domestic violence, forced marriage, rape, torture, child marriage, and assault due to lesbian, gay, bisexual, transgender, and queer/questioning (LGBTQ) status.<br>• Acute FGM/C-related conditions reported: bleeding, infection, shock, broken bones, hospitalization.<br>• Chronic FGM/C-related conditions reported: difficulty with intercourse, pregnancy complications, chronic pain, scarring, difficulty with urination.<br>• Psychological FGM/C-related conditions reported: PTSD, depression, anxiety. |

FGM/C = female genital mutilation/cutting; UTI = urinary tract infection; PTSD = post-traumatic stress disorder
FGM/C type is based on World Health Organization classifications [1] : Type I “Partial or total removal of the clitoris and/or the prepuce (clitoridectomy)”; Type II “Partial or total removal of the clitoris and the labia minora, with or without excision of the labia majora (excision)”; Type III “Narrowing of the vaginal orifice with creation of a covering seal by cutting and appositioning the labia minora and/or the labia majora, with or without excision of the clitoris (infibulation); Type IV “All other harmful procedures to the female genitalia for non-medical purposes, for example: pricking, piercing, incising, scraping and cauterization”

### Characteristics of Included Studies

Among the 40 articles included, 18 were qualitative studies (45%), 16 were quantitative (40%), and 6 were mixed methods (15%) (Figure 2). The first study was published in 1985 [23], and over half (63%) were published since 2014 [9, 24–47]. The sample sizes ranged from 7 to 12,270, with most studies having relatively small sample sizes (under 100). The largest study, by Sudhinaraset et al. [43], with a sample size of 12,270, used a population-based dataset of refugees from Africa, Southeast Asia, Europe/Central Asia, Latin America and the Caribbean, and South Asia who entered California between 2013 and 2017. Studies collected data from participants in a wide range of US locations, with New York-based samples most common [24, 29, 33–35, 37, 41, 45, 48–50]. Most study populations were adults aged 18 or older, with few including individuals under 18 [9, 43, 47, 48, 51]. Study participants largely came from African countries, with Somalia the most common [9, 23, 25–27, 30, 32, 33, 36, 38, 39, 42, 47–49, 52–58]. Four studies included individuals from Middle Eastern and Asian countries [36, 42, 43, 46]. Nine studies included males [26, 32, 34, 36, 42, 44, 51, 55, 57]. Six studies obtained information from healthcare providers [23, 25, 36, 50, 55, 58], and three studies included non-US-based participants [51, 56, 59], but findings from healthcare providers and non-US-based participants are not reported in this review.

**Figure 2.**
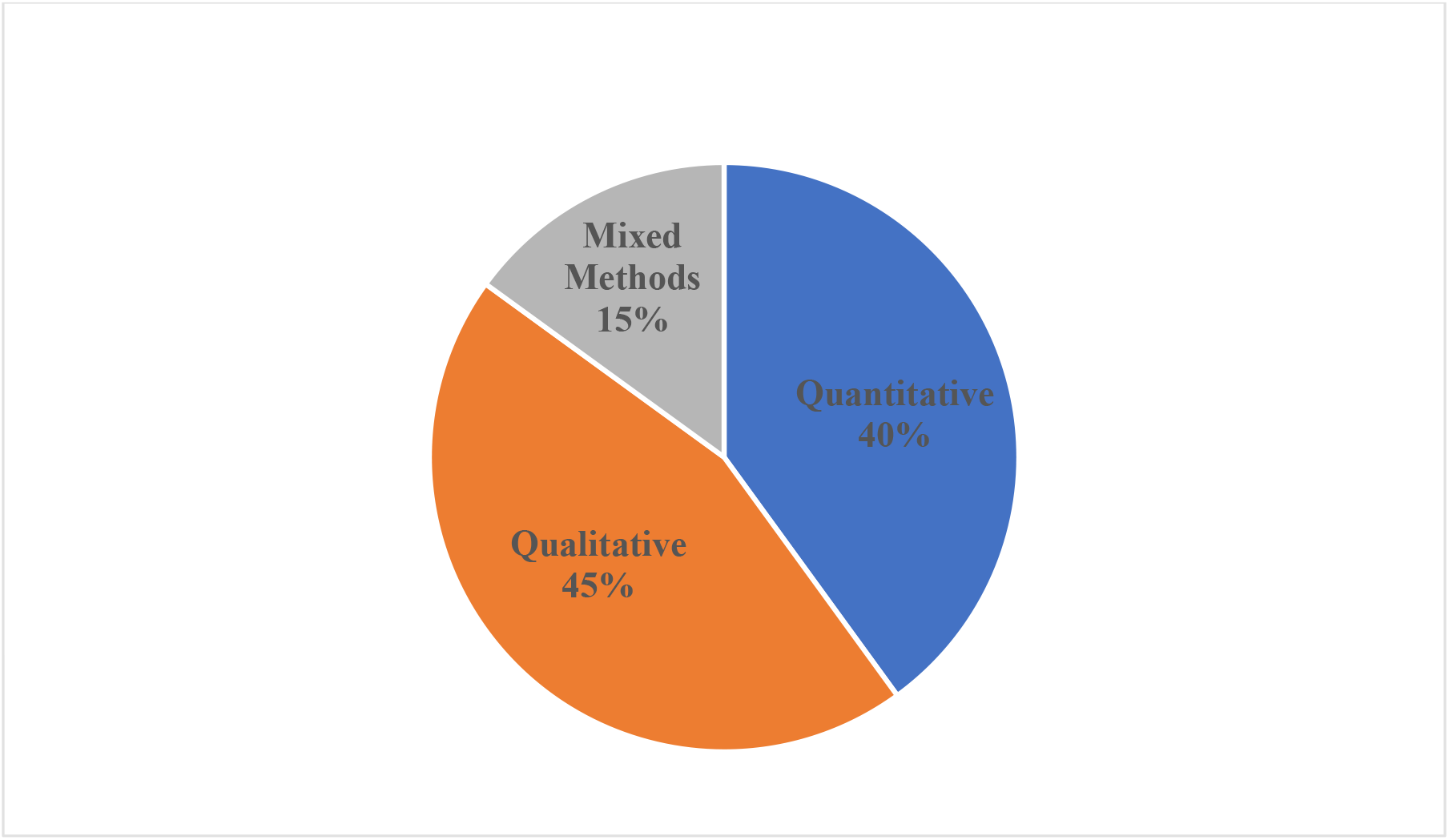
Study design distribution (n = 40)

### Study Findings

#### Attitudes Surrounding FGM/C

Twenty-eight studies examined attitudes about FGM/C [25-34, 36-38, 42, 44, 45, 47-49, 51-56, 58, 60, 61], including support for or opposition to FGM/C; opinions on FGM/C of daughters; FGM/C as a cultural or religious practice; potential harm caused by FGM/C; and FGM/C and attitudes about healthcare experiences and preferences (Table 2).

##### Women’s Attitudes

###### Support for/Opposition to FGM/C

Over half of studies that measured attitudes (n =18) ascertained women’s opinions about whether FGM/C should continue [28-34, 36, 37, 42, 44, 45, 48, 49, 51, 54, 56, 60], with most studies finding substantial opposition to FGM/C. In some studies where women opposed FGM/C, they tended to view it as a form of abuse or violence or as a human rights violation [29, 33, 44]. Some studies found that women had become less supportive of the practice after immigration to the United States [32, 42, 60]; one study comparing more established with recent immigrants found that both groups had negative attitudes about FGM/C [42].

###### FGM/C for daughters

Fourteen studies reported on women’s opinions about FGM/C for daughters, with most studies finding opposition to having daughters undergo FGM/C [27-32, 36, 37, 48, 53, 56, 58, 60, 61]. In at least four of these studies, some women supported having their daughters undergo FGM/C [28, 48, 53, 56]. In one of the studies [48], most women said they would disapprove of their daughters undergoing the most severe form of FGM/C (Type III-infibulation) but would support milder forms of FGM/C (Type I).

Some studies explored women’s thoughts on the role of the community in perceptions related to FGM/C, as well as whether it was important to discuss the issue within the family, particularly with daughters [28, 36, 42, 58, 60]. Two studies found that participants supported discussing FGM/C in communities and recognized the potential influence of communities on changing attitudes and behaviors [36, 42]. However, in at least two studies, women reported having mixed feelings [60] or felt it was unnecessary to discuss FGM/C with their daughters or in their communities [58]. Furthermore, in four studies, women reported pressure from family members in their country of origin or in the United States to have their daughters undergo FGM/C [31, 32, 37, 61].

Vacation cutting was discussed in six studies [31, 36, 37, 42, 44, 53]. Overall, these studies found women were opposed to vacation cutting. In two studies, women discussed fears and feeling pressure from family to take their daughters to their home countries where they would be at risk of undergoing FGM/C without maternal consent [31, 37]. In one study, women discussed having been informed by medical providers that if their daughters had FGM/C after returning from a trip to Africa, there would be legal consequences [37]. In two studies, women reported that they were aware of instances where daughters were sent back for FGM/C [37, 44]. In one study [53], a respondent indicated she had plans to take her daughter to Somalia for FGM/C. In contrast, Shahawy et al. found that participants were unaware of any instances of vacation cutting and felt that it would not be possible because of the legal repercussions [42].

###### Harm/Benefits of FGM/C

Multiple studies assessing opinions on the harms and benefits of FGM/C found that participants reported believing it was a harmful practice for women and girls, both physically and mentally/emotionally [27, 29, 36, 37, 44, 49, 54]. In these studies, women described FGM/C as having adverse effects, with physical health consequences more commonly mentioned than mental/emotional consequences [27, 29, 36, 37, 44, 49, 54]. Several studies explored perceived non-health benefits of FGM/C or thoughts on why the practice is perpetuated [28, 32, 37, 42, 45, 54, 58, 61]. The benefits or rationales cited included supporting cultural practices, finding a husband from the same culture/marriageability, preventing premarital sex, and cleanliness [28, 32, 37, 42, 54, 58, 61]. It is important to note that in most of these studies, women generally opposed the practice but discussed these reasons when asked why the practice persists.

###### FGM/C as a cultural/religious practice

In at least 11 studies, women described their understanding of FGM/C as a religious practice (or requirement), a cultural practice, or both [25, 28, 32, 33, 36, 42, 44, 45, 56, 60, 61]. Across studies, regardless of where respondents came from, they more commonly reported that FGM/C is a cultural rather than religious practice. In one study [32], women reported that before migration they had thought FGM/C, specifically infibulation, was a religious requirement, but since migration they realized this was not the case. In two studies, most women reported believing FGM/C was based in religion [56, 58].

###### FGM/C and attitudes about healthcare experiences and preferences

At least 13 studies reported women’s attitudes about healthcare experiences related to FGM/C [23, 25, 32, 36–38, 47, 50, 52, 53, 56, 58, 61]. Common findings were that women felt that healthcare from US providers was often culturally insensitive and that healthcare providers were unfamiliar with FGM/C and how to treat or deliver women who had undergone it [32, 38, 56, 61]. Women in these studies cited the importance of having a gynecologic provider familiar with and accepting of women with FGM/C. In several studies, respondents reported that they would prefer female healthcare providers [53, 58] and those who had experience or training in FGM/C [38, 61]. Women also reported that they were reluctant to bring up FGM/C with their healthcare providers but welcomed healthcare providers initiating discussions [36, 58]. Ameresekere et al. [52] found that most respondents who had undergone FGM/C reported that healthcare providers never discussed FGM/C with them. In two recent studies, women reported positive attitudes and experiences related to healthcare providers, including a sense of respectful treatment, no discrimination, and feeling comfortable [36, 47].

##### Men’s Attitudes

Eight studies included men and women participants [32, 34, 36, 42, 44, 51, 55, 57] and one study [26] men exclusively. Two of these were excluded from this section, one due to inclusion of only one man [32] and the other because results were not broken out by sex [51]. Studies with men included in the samples tended to be smaller and report fewer results than studies with women.

Men in these seven studies tended to report opposing FGM/C and to state that it should be discontinued. In the studies where such information was ascertained, most men preferred not to have their daughters undergo FGM/C [34, 42, 44]. The Johnson-Agbakwu et al. study [26] showed that participating men were aware of the morbidities FGM/C could cause and the laws against the practice in the United States. The Partnerships for Health study [36] likewise found that men in the study thought FGM/C was harmful to women. The Akinsulure-Smith and Chu study [34] reported that most men in the study had no preference for dating or marrying women with or without FGM/C. Men who did have a preference tended to prefer a woman without FGM/C [34]. Shahawy et al. found that among men in their study, preferences varied and may have changed over time, from preferring a woman with FGM/C to preferring a woman who had not undergone the practice [42]. In two studies [36, 42], most participating men thought it was important that FGM/C be discussed in the community to raise awareness. Finally, Johnson-Agbakwu et al. [26] found that men in their study tended to report that healthcare providers lacked adequate knowledge and training related to FGM/C.

#### Women’s Experiences with FGM/C

Most of the 40 studies included in this review collected data on women’s self-reported FGM/C status, and 12 studies included in the sample only women who experienced FGM/C [23, 31–33, 37, 40, 41, 52, 55–58]. Other aspects of FGM/C assessed included: circumstances surrounding FGM/C; sexual health; gynecologic and obstetric health; experiences in seeking healthcare; mental health; and experiences of violence or trauma in addition to FGM/C (Table 2).

##### FGM/C Status and Circumstances Surrounding FGM/C

A study by Sudhinaraset et al. [43] of female refugees coming to California from any country between 2013 and 2017 had information on FGM/C from 10,132 girls and women and found 2.1% of women ≥15 years of age and 0.6% of girls <15 years of age reported FGM/C. In the same study, among the subset of female African refugee girls (n = 236) and women (n=482) with FGM/C information, 5.5% of girls <15 years of age and 30.1% of those ≥15 years of age reported FGM/C [43]. Additionally, less than 1% of girls and women from each of the other regions (i.e., Southeast Asia, Europe/Central Asia, Latin America and the Caribbean, and South Asia) reported FGM/C (Table 2) [43]. In another study that assessed daughters’ experience of FGM/C among 68 Nigerian mothers, 33% indicated their daughters had FGM/C [28]. Other studies that included women from multiple African countries found differences in the percentage of women with FGM/C by country of origin [24, 29, 35, 45]. For example, in their study of West African women, Chu and Akinsulure-Smith found the percentage with FGM/C differed significantly by country of origin, with ranges from 45% among Sierra Leonean women to 92% among Malian women [29].

At least 16 studies assessed or discussed the type of FGM/C procedure women or their daughters experienced [9, 25, 28–30, 32, 39–41, 46–48, 50, 55–57]. Type III was most often reported, followed by Type II (having flesh removed). Women often reported that they experienced FGM/C during infancy or childhood [31-33, 40, 41, 45, 46, 48, 51, 56]. Those who recounted their FGM/C experience (or the experiences of others) often described it as traumatic, painful, and causing immediate adverse physical health experiences (e.g., infection, bleeding, swelling) [28, 31, 32, 37, 48, 60].

##### Sexual Health

Fifteen studies examined aspects of sexual health among women who experienced FGM/C [9, 23, 27, 29–33, 37, 41, 42, 46, 57, 59, 60]. Women commonly reported a negative impact of FGM/C on sexual relationships, including painful intercourse, lack of sexual pleasure, and lack of desire for intimacy. Differences in sexual health experiences were found according to FGM/C status [29] and FGM/C type [30]. One study of West African immigrants that used the Female Sexual Function Index (FSFI) [62] found no significant differences in overall FSFI scores between women with and without FGM/C (including in the domains of desire, lubrication, orgasm, pain, and satisfaction) [29]. The study did find that women with FGM/C had significantly lower scores on the FSFI arousal domain [29]. Connor et al. [30] found that women with more severe types of FGM/C (Types II or III) were significantly more likely to report painful vaginal intercourse than those with less severe forms (Types I or IV). Nour et al. found that women who undergone defibulation (opening of the infibulation scar [63]) by a medical professional) were more sexually satisfied and that husbands also reported improvements in their sexual life [57].

##### Gynecologic and Obstetric Health

Seventeen studies included information on women’s gynecologic or obstetric health [9, 23, 27–29, 32, 33, 36, 37, 40, 41, 46, 48, 50, 52, 55, 57]. Women with FGM/C commonly reported painful menstruation, vaginal pain, urinary problems, and pelvic pain [9, 23, 27–29, 32, 33, 36, 37, 40, 41, 46, 48, 57]. In two studies [9, 29] that compared women with and without FGM/C, FGM/C emerged as significantly associated with some gynecologic complications. Regarding obstetric experiences and outcomes, women reported experiences with childbirth pain and complications, pregnancy problems, and vaginal tearing [9, 27, 32, 33, 36, 37, 41, 46]. One study that compared women with and without FGM/C found that although there was no direct correlation between FGM/C and type of delivery or overall childbirth experience, women with FGM/C were less likely to report a positive pregnancy experience [36].

##### Experiences in Seeking Healthcare

Seventeen studies explored women’s experiences when seeking or receiving healthcare [9, 23, 25, 27, 32, 36–39, 47, 50, 52, 53, 55, 56, 58, 61]. Negative healthcare experiences included women’s reports of discomfort with treatment, embarrassment by medical staff, lack of communication by healthcare providers about FGM/C, and lack of culturally sensitive care [25, 32, 36, 37, 52, 53, 55, 58, 61]. In some studies, women reported positive experiences, including feeling comfortable or being satisfied with health services [36, 47, 50]. In a recent study, Michlig et al. found that although few women sought healthcare related to their FGM/C, overall, those who did reported positive FGM/C care-related experiences [47]. Some studies compared healthcare seeking among women with and without FGM/C, and with different FGM/C types [9, 36, 39, 61]. Among Somali women, Banke-Thomas et al. [39] found that those with FGM/C were less willing to seek reproductive and maternal care compared with those without FGM/C. There was no association between a woman’s FGM/C status and gaining entry into the health system, accessing a primary care provider, or seeing a specialist [39]. However, when considering FGM/C type, this study found women with more severe FGM/C types (II or III) were less willing to seek care and had more difficulty gaining entry into the health system and accessing a primary care provider compared with women with Type I [39]. Another study of Somali women found those with FGM/C more likely to receive a Pap test and have a place for healthcare compared with women without FGM/C [9]. However, in this same study, among women with FGM/C, the probability of having a designated place for healthcare was lower among women exposed to victimization (home or property looted or burned, abandoned or thrown out by family, abducted, experienced sexual violence, attacked with a weapon, or witnessed a murder) compared to those who were not [9].

##### Mental Health

Eight studies examined FGM/C and mental health outcomes among women, including post-traumatic stress disorder (PTSD), depression, and anxiety [9, 24, 29, 32, 33, 35, 41, 46]. Three found no significant differences in mental health outcomes between women who experienced FGM/C and those who did not [24, 29, 35]. However, Fox and Johnson-Agbakwu [9] found that FGM/C was significantly associated with feelings of depression or flashbacks/nightmares about a traumatic event. Lever et al. [41] found that among 13 women with FGM/C, 92% exhibited anxiety and all exhibited depression. Furthermore, of the 13 women, all 7 who were screened for PTSD had symptoms met the criteria for PTSD [41]. In two qualitative studies [32, 33], women described negative psychological consequences of FGM/C, and McNeely and Christie-de Jong [32] found that 11 of the 12 women in their study reported ongoing depression and anxiety symptoms because of their FGM/C.

##### Trauma or Violence

Five studies explored women’s experience of FGM/C and other forms of trauma or violence [9, 24, 35, 41, 46]. In Lever et al., all women with FGM/C also reported experiencing at least one other type of physical, psychological, or sexual violence, and most reported experiencing multiple types of violence [41]. Akinsulure-Smith and Chu found that a significantly larger proportion of women with FGM/C reported psychological and sexual torture than those without FGM/C [35]. Lastly, Fox and Johnson-Agbakwu found that women with FGM/C who were also exposed to other types of violence (e.g., sexual violence, abduction, witnessing a murder) had more unmet healthcare needs and a higher predicted probability of experiencing problems during pregnancy and gynecological problems than those with FGM/C but not exposed to other types of violence [9].

## DISCUSSION

The studies included in this scoping review represent a growing body of literature exploring FGM/C-related attitudes and experiences of US-resident women and men, many whose lives have been directly or indirectly affected by the practice. In most studies that assessed attitudes surrounding FGM/C, both women and men generally opposed FGM/C. There was opposition to subjecting daughters to the practice, with some studies reporting perceived rejection by family or community members due to conflicting attitudes about FGM/C for daughters, family pressure to have daughters undergo FGM/C, or women’s fears that family members might have girls undergo FGM/C without the mother’s consent [31, 32, 37, 61]. Some studies reported participants’ accounts that coming to the United States influenced them to be less supportive of FGM/C [32, 42, 60].

Physical health problems associated with FGM/C reported in the US literature are similar to those documented in the global literature: gynecological, sexual, and obstetric problems [2, 64, 65]. Severity of the FGM/C procedure is associated with greater health problems, and Type III, the most severe form of FGM/C, was the most common type reported in US studies despite being estimated to account for around 10% of FGM/C cases globally [1]. Over-representation of Type III in US studies may be due to the research emphasis on Somali women, who represent the largest African refugee population in the United States and among whom Type III is common [66, 67].

Women commonly had negative accounts of interactions with health services in the United States and the view that healthcare providers lack cultural sensitivity or understanding of FGM/C [25, 32, 36, 37, 52, 53, 55, 58, 61]. Studies also indicate that negative experiences with the US healthcare system provoked feelings of fear, shame, and humiliation associated with FGM/C. This phenomenon, described as “re-traumatization,” has been cited as a concern for women with FGM/C who migrate to countries where the practice is poorly understood and illegal [68]. Moreover, some women described reluctance to seek healthcare services in the United States because of their own negative perceptions, prior healthcare experiences or due to fear of revealing their daughter’s FGM/C status [37, 39]. Findings from this scoping review indicate the need to provide culturally sensitive and well-informed care for women with FGM/C.

Two other themes that emerged are the adverse mental health effects either directly or indirectly associated with FGM/C and the experience of other types of trauma or violence among many women with FGM/C [9, 32, 33, 35, 41, 46]. Women described traumatic memories of their FGM/C experience, which often occurred during childhood [31]. They also reported ongoing negative effects of FGM/C on their psychological and emotional health [32], including fear and anxiety resulting from their own experience as well as fear for their daughters of undergoing FGM/C without their consent [31, 37]. A number of reviewed studies demonstrated that women with FGM/C had also experienced other types of physical, psychological, or sexual trauma or violence [9, 24, 35, 41, 46]. One found that among women with FGM/C, experiences of other trauma or violence compounded the likelihood of depression and reproductive health problems [9]. These findings indicate the importance of understanding FGM/C in the broader contexts of women’s mental health and other traumatic life experiences rather than treating FGM/C as an isolated physical health condition.

### Strengths and Limitations

The studies included in this review have several strengths. One strength is that they represent varied research methods, including qualitative, quantitative, and mixed methods approaches. A second strength is that the studies had diverse participants, including men, people from multiple countries of origin, and individuals with varied immigration histories. Third, the depth and variety of topics included in the literature provide insight into different aspects of experiences and attitudes surrounding FGM/C, strengthening our understanding of the implications of FGM/C for individuals living in the United States. A fourth strength is that while studies published before 2014 had smaller sample sizes (e.g., ≤50), some more recent research tended to have larger sample sizes, providing greater power for statistical analysis.

The literature also has limitations and notable gaps. One limitation is that although some studies reported prevalence of FGM/C in their study samples, none can be generalized to all migrants in the United States from a specific country of origin or in a US geographic area. In fact, a representative sample that could yield a prevalence estimate for the United States is likely to be unattainable due to information about FGM/C not being systematically collected/reported and difficulty constructing a sampling frame. Studies of US participants from a single country of origin or community may not be comparable to those of people from other countries of origin or communities. A second limitation is that most of the studies relied on convenience and purposive sampling, which can contribute to biases, including coverage, non-response, and/or self-selection bias that limit the interpretation of findings. Third, most included studies relied on self-reported FGM/C status, which may not be a reliable way to document specific FGM/C status or typology [69–71]. Given that FGM/C is illegal in the United States and that it is considered a private and highly personal topic, the study results may have response and/or social desirability biases and may not accurately reflect participants’ experiences or attitudes. Data collected on the illegal practice of families sending children abroad for FGM/C is also subject to response and/or social desirability biases [72]. An understanding of the number of girls at risk for vacation cutting is needed to assess the overall number of US-resident women and girls at risk for FGM/C [13, 73], but no study in this review attempted to assess the number of girls affected or at risk.

The limitations of this scoping review itself should also be noted. As a scoping review, this review synthesizes a wide range of research studies and describes available research, but does not systematically evaluate the evidence to answer a specific research question [17]. The review topics were restricted to women and men’s attitudes and experiences related to FGM/C. Literature on related topics, such as healthcare providers’ attitudes and experiences or perspectives of non-US-based individuals, was not included. Additionally, although we employed a broad search strategy, it is possible that relevant publications were missed, especially in instances where FGM/C was not the primary study topic.

### Recommendations for Future Research

A more comprehensive and accurate understanding of FGM/C in the United States may be possible through research studies that are more inclusive of the ethnic and cultural diversity of the practice. This would expand the data beyond what was most commonly described in this scoping review, which is the most physically recognizable Type III FGM/C (infibulation) (Table 1). In studies where clinical genital exams are not feasible or acceptable to confirm type of FGM/C, validated tools to assist women reporting their FGM/C type more accurately, including visual aids, may have the potential to increase the accuracy of self-reported FGM/C type [74, 75]. Further, medical records were seldom relied upon in the included reviewed studies and could be informative in future FGM/C research. However, efforts would be needed to promote more complete and consistent documentation of FGM/C in medical records.

Future research could place greater emphasis on understanding the cultural or social influences, whether domestic or abroad, that shape changing attitudes toward FGM/C, including the impact of acculturation [76]. It can be beneficial to involve the community in all phases of research, including conceptualization, implementation, interpretation, and dissemination of findings back to the community. Building, nurturing, and sustaining trust with affected communities through Community-Based Participatory Research (CBPR) approaches may be useful [77]. CBPR community advisory boards can seek to engage men, women, elders, youth, faith-based leaders, and ethnic community-based organizations, as they play a crucial role in articulating the communities’ priorities and ensuring research accountability [77, 78].

Future research on populations affected by FGM/C would benefit from applying a health equity lens that considers the larger social, geopolitical, and racialized context of affected communities in the United States, which are often communities of color, migrants, and religious minorities [79, 80]. Taking into consideration structural disadvantages faced by groups affected by FGM/C could help avoid research that stereotypes and “others” immigrants or people of color [76]. Research that explicitly acknowledges and addresses potential biases can improve understanding of attitudes and experiences of FGM/C, increase positive experiences and communication with healthcare providers, reduce delays in seeking care and/or refusal of care for women with FGM/C, and further efforts to prevent the practice in the United States and globally [68, 76].

## CONCLUSION

Although gaps exist, the existing literature provides substantial insight into the FGM/C-related attitudes and experiences among individuals in the United States. Overall, in most studies both women and men in the United States oppose FGM/C, including for their daughters. While the exact number of women and girls in the United States who have undergone FGM/C is unknown, the evidence indicates that women living with FGM/C in the United States have significant health needs and report having had negative healthcare experiences. Findings from this scoping review identified research gaps and the need for better informed and more respectful and knowledgeable physical healthcare for women who have experienced FGM/C. Another research gap noted is the need for more and better mental health services, in particular culturally informed respectful mental health services. Future research can improve measurement of FGM/C by applying a health equity lens and taking into account the cultural and social influences that shape attitudes and experiences related to FGM/C among people affected by FGM/C in the United States.

## Data Availability

Data sharing not applicable to this article as no datasets were generated or analyzed during the current study.

## Disclaimer

The findings and conclusions in this report are those of the author and do not necessarily represent the official position of the Centers for Disease Control and Prevention.

## Acknowledgements

This research was supported in part by an appointment to the Research Participation Program at the Centers for Disease Control and Prevention administered by the Oak Ridge Institute for Science and Education through an interagency agreement between the U.S. Department of Energy and the Centers for Disease Control and Prevention.

## References

1. World Health Organization. Eliminating female genital mutilation: an interagency statement-OHCHR, UNAIDS, UNDP, UNECA, UNESCO, UNFPA, UNHCR, UNICEF, UNIFEM, WHO. World Health Organization; 2008.

2. Berg RC, Underland V, Odgaard-Jensen J, Fretheim A, Vist GE. Effects of female genital cutting on physical health outcomes: a systematic review and meta-analysis. BMJ, Open. 2014;4(11):e006316. doi: 10.1136/bmjopen-2014-006316.

3. Abdalla SM, Galea S. Is female genital mutilation/cutting associated with adverse mental health consequences? A systematic review of the evidence. BMJ Glob Health. 2019;4(4):e001553-e. doi: 10.1136/bmjgh-2019-001553.

4. Shell-Duncan B, Naik R, Feldman-Jacobs C. A State-of-Art-Synthesis of Female Genital Mutilation/Cutting: What Do We Know Now? . Evidence to End FGM/C: Research to Help Women Thrive. New York: Population Council; 2015.

5. Berg RC, Denison E. A tradition in transition: factors perpetuating and hindering the continuance of female genital mutilation/cutting (FGM/C) summarized in a systematic review. Health Care Women Int. 2013;34(10):837–59. doi: 10.1080/07399332.2012.721417.

6. UNICEF. Female Genital Mutilation/Cutting: A Global Concern. New York, NY: UNICEF; 2016.

7. Davis G, Jellins J. Female genital mutilation: Obstetric outcomes in metropolitan Sydney. Aust N Z J Obstet Gynaecol. 2019;59(2):312–6. doi: 10.1111/ajo.12954.

8. Koukkula M, Keskimäki I, Koponen P, Mölsä M, Klemetti R. Female Genital Mutilation/Cutting among Women of Somali and Kurdish Origin in Finland. Birth. 2016;43(3):240–6. doi: 10.1111/birt.12236.

9. Fox KA, Johnson-Agbakwu C. Crime Victimization, Health, and Female Genital Mutilation or Cutting Among Somali Women and Adolescent Girls in the United States, 2017. Am J Public Health. 2020;110(1):112–8.

10. Pub. L. No. 104-208, §645, 110 Stat. 3009. codified at 18 USC §116. United States1996.

11. Pub. L. No. 112–239, §1088, 126 Stat. 1970. United States2013.

12. STRENGTHENING THE OPPOSITION TO FEMALE GENITAL MUTILATION ACT OF 2020. PUBLIC LAW 116–3092021.

13. Goldberg H, Stupp P, Okoroh E, Besera G, Goodman D, Danel I. Female Genital Mutilation/Cutting in the United States: Updated Estimates of Women and Girls at Risk, 2012. Public Health Rep. 2016;131(2):340–7. doi: 10.1177/003335491613100218.

14. Jones WK, Smith J, Kieke B, Jr., Wilcox L. Female genital mutilation. Female circumcision. Who is at risk in the U.S.? Public Health Rep. 1997;112(5):368–77.

15. OASH Press Office: HHS awards $6 Million to Improve Female Genital Cutting-related Health Care Services for Women and Girls in the United States. https://www.hhs.gov/ash/about-ash/news/2016/hhs-awards-6-m-improve-female-genital-cutting-related-health.html (2016.). Accessed November 20 2020.

16. U.S. End FGM/C Network: US End FGM/C Network Resources. https://endfgmnetwork.org/resources/ (2020). Accessed November 20 2020.

17. Arksey H, O’Malley L. Scoping studies: towards a methodological framework. Int J Soc Res Methodol. 2005;8(1):19–32. doi: 10.1080/1364557032000119616.

18. Veritas Health Innovation: Covidence systematic review software. www.covidence.org Accessed 2019.

19. Moher D, Liberati A, Tetzlaff J, Altman DG, Group P. Preferred reporting items for systematic reviews and meta-analyses: the PRISMA statement. PLoS Med. 2009;6(7):e1000097. doi: 10.1371/journal.pmed.1000097.

20. National Heart, Lung, and Blood Institute: Study Quality Assessment Tools: Quality Assessment Tool for Observational Cohort and Cross-Sectional Studies. https://www.nhlbi.nih.gov/health-topics/study-quality-assessment-tools Accessed January 5 2020.

21. Critical Appraisal Skills Programme: CASP Qualitative Checklist. https://casp-uk.net/casp-tools-checklists/ (2018). Accessed January 20 2020.

22. Hong QN, Pluye P, Fàbregues S, Bartlett G, Boardman F, Cargo M, et al. Mixed methods appraisal tool (MMAT), version 2018. 2018;1148552.

23. Shaw E. Female circumcision: perceptions of clients and caregivers. J Am Coll Health. 1985;33(5):193–7. doi: 10.1080/07448481.1985.9939604 10.1080/07448481.1985.9939604.

24. Akinsulure-Smith AM. Exploring female genital cutting among west African immigrants. J Immigr Minor Health. 2014;16(3):559–61.

25. Fawcett L. Somali Refugee Women and Their U.S. Healthcare Providers: Knowledge, Perceptions and Experiences of Childbearing. Ann Arbor: Arizona State University; 2014. p. 348.

26. Johnson-Agbakwu CE, Helm T, Killawi A, Padela AI. Perceptions of obstetrical interventions and female genital cutting: insights of men in a Somali refugee community. Ethn Health. 2014;19(4):440–57.

27. Jacoby SD, Lucarelli M, Musse F, Krishnamurthy A, Salyers V. A Mixed-Methods Study of Immigrant Somali Women’s Health Literacy and Perinatal Experiences in Maine. Journal of Midwifery & Women’s Health. 2015;60(5):593–603.

28. Ukoha DE. Female Genital Mutilation/Circumcision: Culture and Women Sexual Health in Igbo women residing in Dallas-Fort Worth, Texas. Ann Arbor: Walden University; 2015. p. 139.

29. Chu T, Akinsulure-Smith AM. Health Outcomes and Attitudes Toward Female Genital Cutting in a Community-Based Sample of West African Immigrant Women from High-Prevalence Countries in New York City. J Aggress Maltreat Trauma. 2016;25(1):63–83.

30. Connor JJ, Hunt S, Finsaas M, Ciesinski A, Ahmed A, Robinson BB. Sexual Health Care, Sexual Behaviors and Functioning, and Female Genital Cutting: Perspectives From Somali Women Living in the United States. J Sex Res. 2016;53(3):346–59.

31. Kahn S. “You see, one day they cut”: The evolution, expression, and consequences of resistance for women who oppose female genital cutting. J Hum Behav Soc Environ. 2016;26(7-8):622–35. doi: 10.1080/10911359.2016.1238805.

32. McNeely S, Christie-De Jong F. Somali refugees’ perspectives regarding FGM/C in the US. Int J Migr Health Soc Care. 2016;12(3):157–69. doi: 10.1108/IJMHSC-09-2015-0033.

33. Tatah EF. Female Circumcision: A Phenomenological Study of Somalian Immigrants to the United States. Ann Arbor: Walden University; 2016. p. 133.

34. Akinsulure-Smith AM, Chu T. Knowledge and attitudes toward female genital cutting among West African male immigrants in New York City. Health Care Women Int. 2017;38(5):463–77.

35. Akinsulure-Smith AM, Chu T. Exploring Female Genital Cutting Among Survivors of Torture. J Immigr Minor Health. 2017;19(3):769–73.

36. Partnerships for Health. Maine Community and Clinical Perspectives on FGM/C. Augusta, Maine: Partnerships for Health; 2017. p. 1–35.

37. Akinsulure-Smith AM, Chu T, Krivitsky LN. West African Immigrant Perspectives on Female Genital Cutting: Experiences, Attitudes, and Implications for Mental Health Service Providers. J Int Migr Integr. 2018;19(2):259–76. doi: 10.1007/s12134-018-0544-6.

38. Mehta PK, Saia K, Mody D, Crosby SS, Raj A, Maru S, et al. Learning from UJAMBO: Perspectives on Gynecologic Care in African Immigrant and Refugee Women in Boston, Massachusetts. J Immigr Minor Health. 2018;20(2):380–7.

39. Banke-Thomas A, Agbemenu K, Johnson-Agbakwu C. Factors Associated with Access to Maternal and Reproductive Health Care among Somali Refugee Women Resettled in Ohio, United States: A Cross-Sectional Survey. J Immigr Minor Health. 2019;21(5):946–53. doi: 10.1007/s10903-018-0824-4.

40. Geynisman-Tan J, Milewski A, Dahl C, Collins S, Mueller M, Kenton K, et al. Lower Urinary Tract Symptoms in Women With Female Genital Mutilation. Female Pelvic Med Reconstr Surg. 2019;25(2):157–60.

41. Lever H, Ottenheimer D, Teysir J, Singer E, Atkinson HG. Depression, Anxiety, Post-traumatic Stress Disorder and a History of Pervasive Gender-Based Violence Among Women Asylum Seekers Who Have Undergone Female Genital Mutilation/Cutting: A Retrospective Case Review. J Immigr Minor Health. 2019;21(3):483–9.

42. Shahawy S, Amanuel H, Nour NM. Perspectives on female genital cutting among immigrant women and men in Boston. Soc Sci Med. 2019;220:331–9.

43. Sudhinaraset M, Cabanting N, Ramos M. The health profile of newly-arrived refugee women and girls and the role of region of origin: using a population-based dataset in California between 2013 and 2017. Int J Equity Health. 2019;18(1):158.

44. Ukachukwu UE. Perceptions of Female Genital Cutting among Nigerian Immigrants in Portland, Oregon. Ann Arbor: Walden University; 2019. p. 128.

45. Martell S, Schoenholz R, Chen VH, Jun I, Bach SC, Ades V. Perceptions of Female Genital Mutilation/Cutting (FGM/C) among Asylum Seekers in New York City. J Immigr Minor Health. 2020;13:13.

46. Wikholm K, Mishori R, Ottenheimer D, Korostyshevskiy V, Reingold R, Wikholm C, et al. Female Genital Mutilation/Cutting as Grounds for Asylum Requests in the US: An Analysis of More than 100 Cases. J Immigr Minor Health. 2020;22(4):675–81. doi: 10.1007/s10903-020-00994-8.

47. Michlig G, Warren N, Berhe M, Johnson-Agbakwu C. Female genital mutilation/cutting among somali women in the U.S. state of arizona: Evidence of treatment access, health service use and care experiences. Int J Environ Res Public Health. 2021;18(7).

48. Arbesman M, Kahler L, Buck GM. Assessment of the impact of female circumcision on the gynecological, genitourinary and obstetrical health problems of women from Somalia: literature review and case series. Women Health. 1993;20(3):27–42.

49. Brown E, Carroll J, Fogarty C, Holt C. “They get a C-section…they gonna die”: Somali women’s fears of obstetrical interventions in the United States. J Transcult Nurs. 2010;21(3):220–7.

50. Eyega Z, Conneely E. Facts and fiction regarding female circumcision/female genital mutilation: a pilot study in New York City. J Am Med Womens Assoc. 1997;52(4):174–8, 87.

51. Anuforo PO, Oyedele L, Pacquiao DF. Comparative study of meanings, beliefs, and practices of female circumcision among three Nigerian tribes in the United States and Nigeria. J Transcult Nurs. 2004;15(2):103–13.

52. Ameresekere M, Borg R, Frederick J, Vragovic O, Saia K, Raj A. Somali immigrant women’s perceptions of cesarean delivery and patient-provider communication surrounding female circumcision and childbirth in the USA. Int J Gynaecol Obstet. 2011;115(3):227–30.

53. Beine K, Fullerton J, Palinkas L, Anders B. Conceptions of prenatal care among Somali women in San Diego. J Nurse Midwifery. 1995;40(4):376–81. doi: 10.1016/0091-2182(95)00024-e.

54. Deason L, Githiora R. African Immigrant Women in the United States: Perceptions on Female Circumcision and Policies that Outlaw the Practice. 2013.

55. Horowitz CR, Jackson JC. Female “circumcision”: African women confront American medicine. J Gen Intern Med. 1997;12(8):491–9.

56. Morris R. The culture of female circumcision. Adv Nurs Sci. 1996;19(2):43–53.

57. Nour NM, Michels KB, Bryant AE. Defibulation to treat female genital cutting: Effect on symptoms and sexual function. Obstet Gynecol. 2006;108(1):55–60.

58. Upvall MJ, Mohammed K, Dodge PD. Perspectives of Somali Bantu refugee women living with circumcision in the United States: a focus group approach. Int J Nurs Stud. 2009;46(3):360–8.

59. Nyairo CB. Female Genital Mutilation and Marital Satisfaction among Kenyan Females. Ann Arbor: Loma Linda University; 2013. p. 93.

60. Dahlen UM. Female Genital Cutting: Phenomenological Interviews on the Ethiopian Immigrant Mothers’ Experience. Ann Arbor: Regent University; 2011. p. 318.

61. Gali MA. Female circumcision: A transcultural study of attitudes, identity and reproductive health of East African immigrants. Ann Arbor: The Wright Institute; 1997. p. 132.

62. Rosen R, Brown C, Heiman J, Leiblum S, Meston C, Shabsigh R, et al. The Female Sexual Function Index (FSFI): a multidimensional self-report instrument for the assessment of female sexual function. J Sex Marital Ther. 2000;26(2):191–208. doi: 10.1080/009262300278597.

63. Abdulcadir J, Marras S, Catania L, Abdulcadir O, Petignat P. Defibulation: A Visual Reference and Learning Tool. J Sex Med. 2018;15(4):601–11. doi: 10.1016/j.jsxm.2018.01.010.

64. Berg RC, Underland V. Gynecological Consequences of Female Genital Mutilation/Cutting (FGM/C). NIPH Systematic Reviews: Executive Summaries. Oslo, Norway2014.

65. Berg RC, Odgaard-Jensen J, Fretheim A, Underland V, Vist G. An updated systematic review and meta-analysis of the obstetric consequences of female genital mutilation/cutting. Obstet Gynecol Int. 2014;2014:542859-. doi: 10.1155/2014/542859.

66. Mossad N, Baugh R. Refugees and Asylees: 2016. Washington, DC: Department of Homeland Security, Office of Immigration Statistics; 2018.

67. UNICEF. Female genital mutilation/cutting: a statistical overview and exploration of the dynamics of change. New York, NY: UNICEF; 2013.

68. Hamid A, Grace KT, Warren N. A Meta-Synthesis of the Birth Experiences of African Immigrant Women Affected by Female Genital Cutting. Journal of Midwifery & Women’s Health. 2018;63(2):185–95. doi: 10.1111/jmwh.12708.

69. Bjalkander O, Grant DS, Berggren V, Bathija H, Almroth L. Female genital mutilation in sierra leone: forms, reliability of reported status, and accuracy of related demographic and health survey questions. Obstet Gynecol Int. 2013;2013:680926. doi: 10.1155/2013/680926.

70. Elmusharaf S, Elhadi N, Almroth L. Reliability of self reported form of female genital mutilation and WHO classification: cross sectional study. BMJ. 2006;333(7559):124. doi: 10.1136/bmj.38873.649074.55.

71. Snow RC, Slanger TE, Okonofua FE, Oronsaye F, Wacker J. Female genital cutting in southern urban and peri-urban Nigeria: self-reported validity, social determinants and secular decline. Trop Med Int Health. 2002;7(1):91–100. doi: 10.1046/j.1365-3156.2002.00829.x.

72. Pyati A, De Palma C. Female Genital Mutilation in the United States Protecting Girls and Women in the U.S. from FGM and Vacation Cutting. New York, NY: Sancturary for Families; 2013. p. 1–37.

73. Atkinson HG, Ottenheimer D, Mishori R. Public Health Research Priorities to Address Female Genital Mutilation or Cutting in the United States. Am J Public Health. 2019;109(11):1523–7. doi: 10.2105/AJPH.2019.305259.

74. Abdulcadir J, Dewaele R, Firmenich N, Remuinan J, Petignat P, Botsikas D, et al. In Vivo Imaging–Based 3-Dimensional Pelvic Prototype Models to Improve Education Regarding Sexual Anatomy and Physiology. J Sex Med. 2020;17(9):1590–602. doi: https://doi.org/ 10.1016/j.jsxm.2020.05.025.

75. Wahlberg A, Johnsdotter S, Selling KE, Källestål C, Essén B. Baseline data from a planned RCT on attitudes to female genital cutting after migration: when are interventions justified? BMJ, Open. 2017;7(8):e017506. doi: 10.1136/bmjopen-2017-017506.

76. Johnson-Agbakwu CE, Manin E. Sculptors of African Women’s Bodies: Forces Reshaping the Embodiment of Female Genital Cutting in the West. Arch Sex Behav. 2020. doi: 10.1007/s10508-020-01710-1.

77. Johnson CE, Ali SA, Shipp MP. Building community-based participatory research partnerships with a Somali refugee community. Am J Prev Med. 2009;37(6 Suppl 1):S230–6. doi: 10.1016/j.amepre.2009.09.036.

78. Njie-Carr VPS, Sabri B, Messing JT, Ward-Lasher A, Johnson-Agbakwu CE, McKinley C, et al. Methodological and Ethical Considerations in Research With Immigrant and Refugee Survivors of Intimate Partner Violence. J Interpers Violence. 2019:886260519877951. doi: 10.1177/0886260519877951.

79. Johnson-Agbakwu CE, Ali NS, Oxford CM, Wingo S, Manin E, Coonrod DV. Racism, COVID-19, and Health Inequity in the USA: a Call to Action. J Racial Ethn Health Disparities. 2020. doi: 10.1007/s40615-020-00928-y.

80. Young J. Somali American Adolescent Girls and Women-A Hidden Refugee Population With Barriers to Health. Am J Public Health. 2020;110(1):18–9. doi: 10.2105/ajph.2019.305455.<colcnt=1>

